# Repetition in Social Contact Interactions: Implications in Modelling the Transmission of Respiratory Infectious Diseases in Pre-pandemic & Pandemic Settings

**DOI:** 10.1101/2024.02.09.24302560

**Authors:** Neilshan Loedy, Jacco Wallinga, Niel Hens, Andrea Torneri

## Abstract

The spread of viral respiratory infections is intricately linked to human interactions, and this relationship can be characterised and modelled using social contact data. However, many analyses tend to overlook the recurrent nature of these contacts. To bridge this gap, we undertake the task of describing individuals’ contact patterns over time, by characterising the interactions made with distinct individuals during a week. Moreover, we gauge the implications of this temporal reconstruction on disease transmission by juxtaposing it with the assumption of random mixing over time. This involves the development of an age-structured individual-based model, utilising social contact data from a pre-pandemic scenario (the POLYMOD study) and a pandemic setting (the Belgian CoMix study), respectively. We found that accounting for the frequency of contacts impacts the number of new, distinct, contacts, revealing a lower total count than a naive approach, where contact repetition is neglected. As a consequence, failing to account for the repetition of contacts can result in an underestimation of the transmission probability given a contact, potentially leading to inaccurate conclusions when using mathematical models for disease control. We therefore underscore the necessity of acknowledging the longitudinal nature of contacts when formulating effective public health strategies.

## 1. Background

The propagation of infectious diseases is characterised by various modes of transmission. Transfer of pathogenic agents can occur through contact with fomites, vectors (e.g., insects or animals), airborne aerosols, droplets, or direct physical contact [1]. Notably, viral respiratory infections, primarily transmitted during social interactions via droplets, aerosols and physical contact continue to be major contributors to global morbidity and mortality [2]. Recognising the significance of information on close interactions is pivotal for elucidating disease transmission [3]; thus, social contact data is often employed as a valuable proxy for identifying transmission routes. This information has primarily been collected through diary-based social contact surveys in various countries [4]. An important study is the groundbreaking POLYMOD study which is the first large-scale study which gathered data from eight countries in the European Union [5]. Numerous studies similar in setup have been conducted in different countries in both pre-pandemic and pandemic periods [4]. The CoMix study is the largest study across Europe gathering social contact data aiming to assess changes in contact behaviour during the pandemic [2,6–11]. The information from these surveys has proven invaluable in informing transmission dynamics within mathematical models including network models [12], agent-based models [13], and compartmental models [14–16].

In contrast to our everyday experience, many mathematical models commonly assume a homogeneous mixing [17]. However, research demonstrates that mixing patterns exhibit heterogeneity influenced by factors like age, location, household sizes, intimacy levels, and frequency of contact [5]. Incorporating this heterogeneity in mathematical models is therefore essential to accurately describe transmission dynamics. Various techniques have been developed to achieve this by leveraging social contact data [18]. These methods often involve the use of contact matrices, which consider different contact rates for different subgroups of individuals within a structured population. The contact rate is typically calculated using the average number of contacts reported on a daily basis [19], implicitly assuming that the probability of contacting the same person more than once is negligible and the time between successive contacts within a given age bracket follows an exponential distribution. This contrasts with our daily experience, where we, e.g., frequently interact with household members, a significant aspect of mixing patterns that is not entirely accounted for by modelling methods relying on contact matrices.

Eames et al, (2008) and Smieszek et al, (2009) have demonstrated the importance of considering contact repetition in representing disease transmission [20,21]. Their studies reveal that contact repetition can limit the spread of infection, significantly impacting epidemiological measures when the number of contacts and probability of transmissions are low, as opposed to a random mixing model. However, these studies primarily focused on the theoretical framework and the exploration of repetition in mixing patterns using data has not been fully realised. More recently, Pung et al, (2023) also pointed out the importance of considering the temporal components of contact patterns, highlighting the implication of contact retention by temporally reconstructing dynamic contact networks using data collected via close-proximity sensors [22]. While the use of sensor data provides advantages, the analysis conducted is reliant on information from specific locations and may not comprehensively capture real-life contacts typical in open populations.
Building on previous research, our objective is to reconstruct temporal contact patterns of subgroups within the population and calculate the age-specific number of distinct contacts over a one-week period. We use social contact data obtained from contact diary surveys [2,5], exploring various age groups and different countries to elucidate contact patterns in various settings. Furthermore, we also consider the effect of non-pharmaceutical interventions, delineating how these temporal patterns fluctuated during the COVID-19 pandemic. With the reconstructed patterns, we developed an individual-based model to assess the influence of considering contact repetition, obtained from temporal contact reconstruction.

## 2. Methods

### 2.1 Temporal reconstructions of individuals’ contact patterns

Drawing from data obtained through the POLYMOD and Belgian CoMix studies [2,5], we reconstructed individuals’ contact patterns over a, without loss of generality, one-week period to replicate the social contact dynamics of each individual. In our main analysis, ‘contact’ is defined as physical interactions, including both conversational and non-conversational encounters involving skin-to-skin touching. This approach provides a more nuanced representation of age-specific seroprevalence patterns observed in airborne infections compared with non-physical interactions [12]. Using information from the POLYMOD study, we first scrutinised mixing patterns within a pre-pandemic setting. Subsequently, we consider a pandemic scenario relying on the data collected within the Belgian CoMix study. In the latter case, we addressed the under-reporting issues by expanding upon the method used by Loedy et al, (2023). This correction involved adjusting the reported number of contacts to accommodate under-reporting and redistributing contacts based on the reported frequency for each individual. The choice of the Belgian CoMix study was motivated by its extensive data collection period, allowing us to observe the impact of interventions on reported contacts over an extended timeframe. Notably, the initial eight waves of the Belgian CoMix study lacked information on participants younger than 9 years old, prompting our focus on the later waves. Details regarding the utilised datasets are presented in the Supplementary Materials.

We divided the population into four age categories: Children (0-12 years), Teens (13-18 years), Adults (19-65 years), and the Elderly (66+ years) and reconstructed temporal contact patterns for each of these subgroups. We did so by considering two approaches, a so-called naive approach, where only the number of contacts reported on a daily basis is considered and no frequency is accounted for, and the frequency-based approach. In the latter case, we focused on using the reported frequency of contacts, which indicates how often participants interacted with those they contacted, to reconstruct individuals’ contact patterns over a week. We categorised these interactions into two groups: “daily contacts” (reported as occurring daily in the survey), involving interactions with the same individuals every day, and “non-daily contacts” (reported as weekly, monthly, a few times a year, or for the first time in the survey) - encompassing interactions with different individuals throughout the week. Subsequently, we redistribute the number of distinct contacts over *d* = 7 days using a multinomial distribution, where *d* is an index over days in the week (Monday, Tuesday, …, Sunday). The probabilities for this redistribution are derived from the data, acknowledging that individuals may have varying numbers of distinct contacts each day throughout the week. To accommodate data variability, we undertake the reconstruction of contacts for each individual *i*, by randomly sampling a value from the Generalised Poisson distribution, allowing us to describe under- and over-dispersion in the data, to represent their everyday contacts [23,24]. We define *k* as an index representing the frequency of contact, partitioned into five categories (daily, weekly, monthly, a few times a year, first time) and *a* serves as an index for age groups (0-12, 13-18, 19-65, 66+). Let μ_*a,k*_ and σ_*a,k*_^2^ be the mean and variance, respectively, for age group *a* with contact frequency *k*. Mathematically, the frequency-based approach can be expressed as

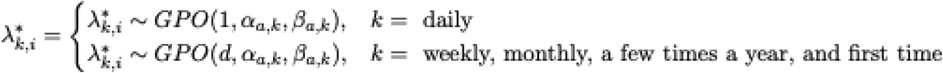

with 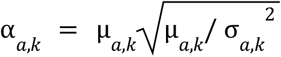 and 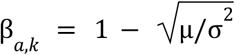. An example showing the outcome of the frequency-based reconstruction is presented in Table S1. Relying on this framework and the considered approaches to reconstruct contact patterns, the total number of distinct contacts in a week is calculated as 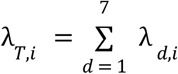 and 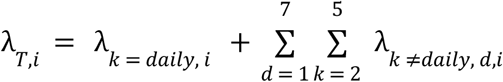 for the naive and the frequency-based approach, respectively.

### 2.2 Epidemic simulation and scenarios

To investigate the impact of temporal contact patterns on transmission dynamics, we developed a discrete-time age-structured stochastic individual-based model in which contact patterns are implemented using either the naive or the frequency-based approaches described previously. In both cases, epidemics start with seeding one index case in a closed and fully susceptible population and will persist until no infected individuals are present [25]. We simulated contact interactions for each infected individual, and we conducted a Bernoulli trial with probability *q* to assess whether the specific interaction led to a transmission event. Within both approaches, such a probability is constant and of the same value among the different age groups, but it is set to describe a different value of the reproduction number according to the social contact hypothesis [19]. In the naive approach, at each time step, every infected individual comes into contact with *c*_*i*_ individuals drawn randomly from the population, setting the within and between mixing rate among the different age groups from the observed data. In the frequency-based approach, for each individual *i*, we randomly selected daily contacts, with whom *i* would have an interaction every day, and non-daily contacts, sampling different individuals who were not part of the daily interactions.

We consider an S-I-R type of dynamics [26], assuming the infectious period to be geometrically distributed *I* ∼ *Geo*(γ), with an average of 7 days. We further set the values of the reproduction number (*R*_0_) equal to: *R*_0_= 1.3, to represent diseases with the spreading potential of a seasonal influenza-like illness [27], and *R*_0_ = 3.3, reflecting diseases with the spreading potential of a COVID-19-like infection [28]. Fixing the population size at *N* = 5000, we ran 3000 simulations for each considered scenario. From the simulated epidemics, we only considered the non-extinct outbreaks, defined as the outbreaks where at least 10% of the population was eventually infected, and we calculated and reported attack rates, time to peak, peak incidence, and epidemic durations as epidemiological summary measures.
The simulations were run for all eight countries participating in the POLYMOD study and 3 different waves of the CoMix study for Belgium. These three different waves are wave 19 (mandatory telework and mask-wearing, curfew, part-time distance learning in schools, maximum 1 (2 if one lives alone) close contact, and full closure (except take away) for restaurants, cafes, and bars), wave 27 (mandatory mask-wearing, compulsory telework, maximum 8 people per table when dining in restaurants, cafes, and bars), and wave 43 (mandatory mask-wearing and COVID safe ticket) which represent three different stages in the COVID-19 pandemic intervention [2].

## 3. Results

### 3.1. Pre-pandemic setting

#### 3.1.1. Contact repetition

The proportions of daily and non-daily physical contacts for the pre- and pandemic scenarios are presented in Figure 1 and Figure S2, respectively. In the former case, we first noticed that age affects the frequency of contact. In most of the countries considered, children and teenagers reported more daily than non-daily contacts, while adults and the elderly reported more non-daily contacts.

Following the methodology presented in Section 2.1., we calculated the total number of distinct contacts using the naive and frequency-based approach for different European countries. Failing to account for repetition leads to overestimating the total number of different contacts occurring in one week (Figure 2). This pattern holds across different age groups and countries. For the Elderly, the effect is considerably smaller than other age categories as the ratio of frequency-based and naive distinct contacts is closer to 1. We further notice that the relative difference between the total number of contacts made by specific age groups varies depending on the approach considered to reconstruct contact patterns, possibly affecting the contribution of such age groups in transmission (Figure S3).

**Figure 1.**
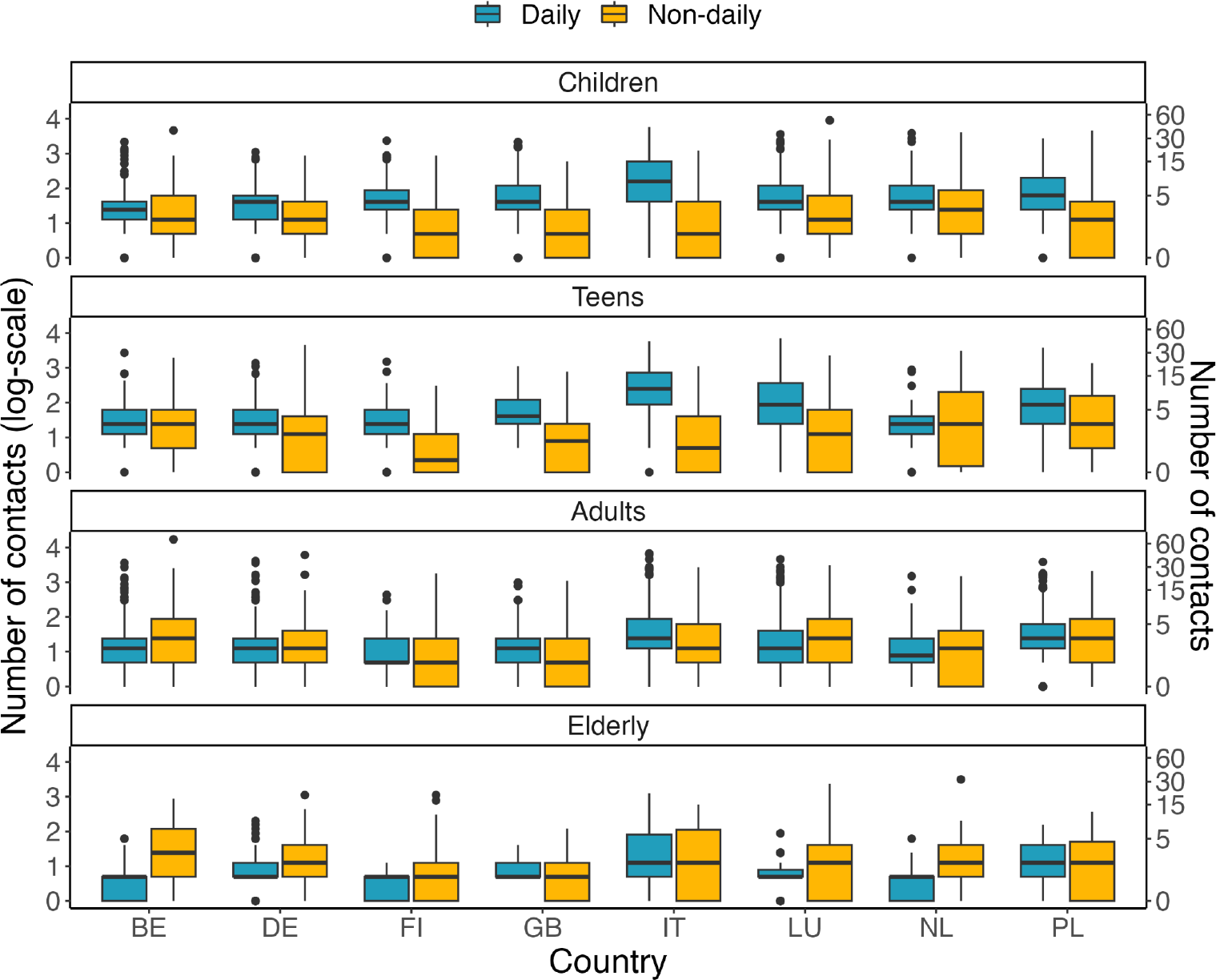
The distribution of the total number of daily and non-daily physical contacts from the POLYMOD study.

**Figure 2.**
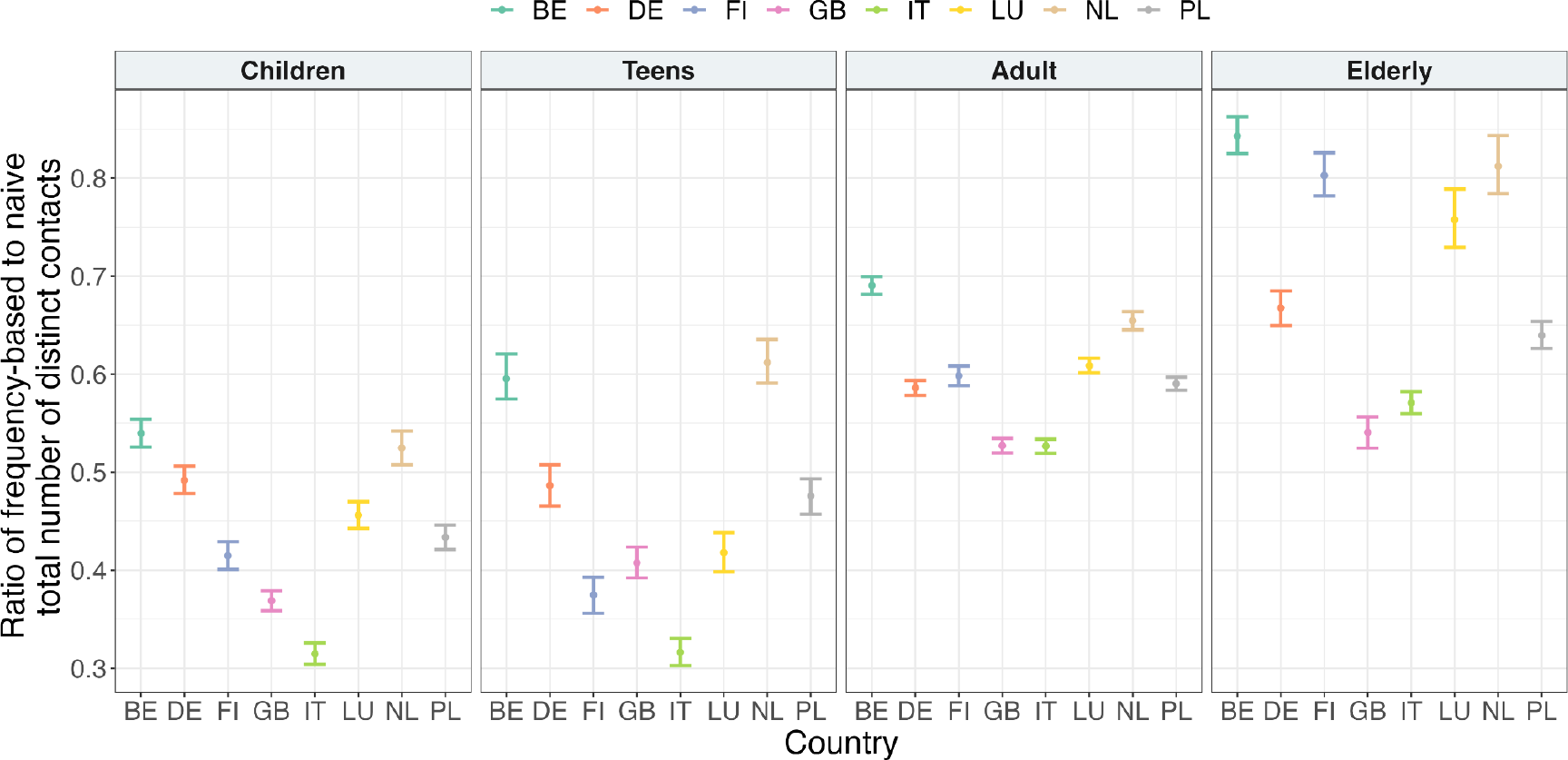
The ratio between frequency-based and naively calculated distinct contacts over a week in a pre-pandemic scenario, together with its 95% Confidence Interval obtained by a non-parametric bootstrap for Children, Teens, Adults, and the Elderly in different countries.

#### 3.1.2. Outbreak characteristics

Results of the simulation study suggest that epidemics are affected by the contact pattern considered over time. Consistently, the attack rate is observed to be higher when the naive reconstruction is assumed, especially when the reproduction number is lower (Figure 3 and Figure S4). Furthermore, the frequency-based approach consistently results in a lower peak incidence (Figure S5) and a longer time to peak. Regarding the overall epidemic duration, the difference is not apparent between the frequency-based and naive approaches for influenza-like illness, but a slight discrepancy is observed for COVID-19-like illness, with the former leading to epidemics with longer duration. In addition, a higher extinction rate is also observed when the frequency-based approach is considered (Table S2).

**Figure 3.**
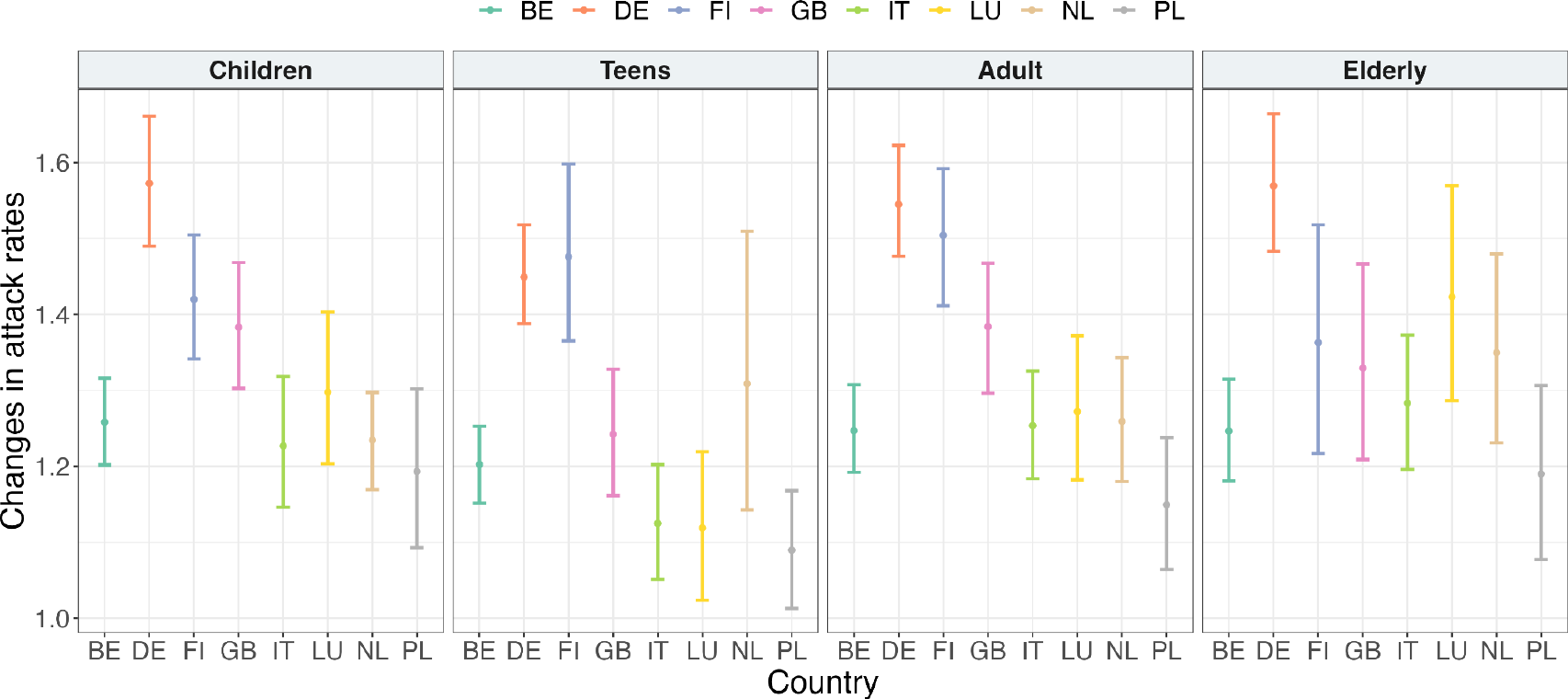
Changes in epidemic attack rates in influenza-like illness when simulating epidemics using a frequency-based approach compared to a naive approach during a pre-pandemic scenario, along with non-parametric bootstrapping for 95% confidence intervals.

### 3.2. Pandemic setting

#### 3.2.1. Contact repetition

In addition to investigating the impact of temporal reconstruction across different countries in Europe, we further describe how such contact profiles vary during a pandemic setting, by using the Belgian CoMix data as a case study. A different behaviour can be seen during the pandemic scenario, where for all age categories, the majority of reported contacts are daily contacts (Figure S2). The ratio of the number of distinct contacts between the frequency-based and naive approaches during the pandemic is greater than that of the pre-pandemic period with the decrease varying across age categories, with more variability shown in the elderly (Figure S6). While for adults and the elderly, the total number of distinct contacts seems to be stable over the different ways, a higher variation is observed for children and teenagers (Figure S7). In addition, for children and teenagers, we observed a higher difference between distinct contacts reconstructed with the naive and the frequency-based approach. As expected, we observed that the number of distinct contacts is greatly reduced when comparing it with the number of contacts during the pre-pandemic scenario. This holds true for all and only physical contacts.

#### 3.2.2 Outbreak characteristics

Considering the same respiratory infections assumed previously, we simulate epidemics during a pandemic scenario. Calculating attack rates using the frequency-based approach results in an underestimation, ranging from approximately 25-45% in influenza-like scenarios to about 60-85% in COVID-19-like scenarios when compared to the naive approach (Figure S8). This effect is also more pronounced in survey waves with stricter interventions (wave 19 compared to wave 43). When evaluating other epidemiological measures, we observed a lower peak incidence when simulating the epidemic with a frequency-based approach in the pandemic scenario (Figure S9), while the impact on the epidemic duration and time to peak was less pronounced in the pre-pandemic scenario and extended over a longer period in the pandemic scenario. Lastly, extinction rates depend on the reproduction number assumed but are always computed to be higher for the epidemics simulated with the naive approach, showing more pronounced relative differences in the pandemic scenario (Table S3).

### 3.3. Influential factors

We examined mixing patterns with respect to intimacy level and found more pronounced differences in the total number of distinct contacts when specifically considering physical interactions, compared to analysing all contacts (Figure S10). Furthermore, we consider the location where contacts occur by comparing interactions that take place within and outside the household setting. It can be seen that during the pre-pandemic period, the majority of contacts occurred outside the household (Figure S11 and Figure S12), while during the pandemic, household interactions were the main components of mixing patterns. In particular, we observed that Children and Teenagers show high variability in terms of the number of contacts outside the home during the pandemic, possibly caused by the interventions in place. We examined the impact of temporal reconstruction on transmission dynamics for different lengths of the infectious period, observing as expected that the influence is less pronounced for shorter infection periods and becomes more significant for longer durations (Table S4).

## 4. Discussion

Social interactions are quintessential for characterising and describing the transmission of respiratory infectious diseases, constituting a key element in modelling transmission dynamics. Population-level epidemic models often neglect the temporal component of human contact behaviour, treating interactions occurring on a specific day as independent from those on other days. In this study, we reconstructed contact interactions over the course of one week and investigated the implications of such a reconstruction in modelling the transmission of respiratory diseases. In particular, we considered two epidemiological settings in which human interactions have been shown to differ widely, i.e., during a pre-pandemic and a pandemic stage.
Our findings align with established evidence from other studies regarding the heterogeneity of contact behaviour, such as age, intimacy level, contact location, and country [5,12,19,29,30]. We further demonstrated a high proportion of contact repetition, emphasising its influence on contact patterns over time. Considering the temporal aspect of contact interactions, especially in epidemiological contexts characterised by a high frequency of contact repetition, such as schools and workplaces, could provide valuable insights to develop intervention strategies in these locations. Furthermore, we identified an increase in the proportion of daily contacts, along with a decrease in the total number of contacts during the COVID-19 pandemic, likely caused by the non-pharmaceutical interventions in place at the time [9]. This finding can provide useful insights into setting population-level interventions for future epidemic or pandemic threats, and further underlines the importance of collecting comprehensive contact data in various settings, ensuring a better understanding of different human interactions.

One of the key elements that characterise super-spreading events is a particularly high number of interactions [31,32]. In the attempt to gain insights into the drivers of super-spreading events, one could reconstruct the number of distinct contacts made by specific groups of individuals in specific locations. Our study highlights the significance of accounting for the longitudinal aspect of contact behaviour, specifically addressing the potential repetition of contacts. Linked with viral shedding kinetics, environmental factors, and other contact characteristics (e.g., duration) [28], the total number of distinct contacts provides crucial insights for identifying the circumstances under which superspreading events can take place.

To unravel the implications of accounting for temporal contact patterns in modelling, we simulated epidemics by implementing both a contact structure that accounts for repetition, i.e., frequency-based approach, and a contact structure in which contact behaviour is considered independent during consecutive days, i.e., naive approach. The results indicate that omitting the temporal component of contact behaviour might lead to an underestimation of the transmission parameter [17], and this becomes particularly noticeable with an increase in contact repetitions, typical of the pandemic period. Moreover, we investigated other outbreak characteristics (e.g., time to peak, peak incidence, and epidemic durations), showing an impact of accounting for the longitudinal aspect of contact interactions. We acknowledge that the reproduction number may differ between the two approaches, potentially influencing the observed differences. While outbreak characteristics are shown to depend on the temporal nature of contact interaction, we observed that accounting for temporal components has a lower impact on infections with high transmissibility, in line with what has been previously observed by Smieszek et al (2009). Therefore, it is crucial to acknowledge that, apart from contact patterns and transmission probability, other characteristics (e.g., infectious period distribution) also play a role in shaping the epidemiological outcome of an epidemic [33].

Social contacts exhibit greater complexity than the model assumptions. Specifically, our assumption of daily contact, where an individual interacts every day with the same person, may lead to an overestimation of repetition. In contrast, our assumption of non-daily contact, occurring each time with a different individual, may underestimate the impact of repetition. However, we focused on a time frame of one week, where such assumptions are supposed to have a limited effect on the overall results. Additionally, we only accounted for contact reciprocity at the population level rather than the individual level (e.g., if *i* is a daily contact of *j*, this does not imply that *j* is a daily contact of *i*) [17]. Further investigations are needed to assess whether individual reciprocity may have additional effects. Furthermore, we restricted our analysis to the stochastic SIR epidemic model. While it is possible to extend this to more complex models, we expect the general findings established in this paper will similarly apply to other models and modelling frameworks [21]. Finally, CoMix is a longitudinal survey where participants may experience fatigue as the survey progresses [2]. When correcting for the under-reporting, we implicitly assume that the impact of fatigue is consistent for both daily and non-daily contacts. However, it can be argued that this effect might vary across different contact frequencies. One approach could be to ask about daily contacts once and only ask about non-daily contacts in later survey waves, as a way to minimise under-reporting while completing the survey.

## 5. Conclusions

This study describes the frequency of social interactions responsible for the transmission of respiratory infectious diseases, in several EU countries in a pre-pandemic setting and during the COVID-19 pandemic in Belgium. Our findings underscore the importance of considering longitudinal aspects of social contact data when assessing the total number of infections attributable to an infectious case. Additionally, we emphasise the significance of investigating the temporal component in population-level models for the spread of infectious diseases. Failure to account for the repetition of contacts may result in an underestimation of the transmission potential of infectious diseases. Moreover, our analysis reveals that mixing patterns are contingent upon individual characteristics, such as age, and specific locations, including households. These insights provide valuable guidance for shaping both local and global intervention measures to limit the spread of respiratory infections.

## Data Availability

The datasets analysed during the current study are available in the Socrates tool (www.socialcontactdata.org/data), as well as Zenodo-based repository (https://zenodo.org/record/7086043#.Y0Zt9C8RrmE) .

## Data accessibility

The datasets analysed during the current study are available in the Socrates tool (www.socialcontactdata.org/data), as well as Zenodo-based repository (https://zenodo.org/record/7086043#.Y0Zt9C8RrmE) . The source code for this project is available on GitHub at (https://github.com/neilshanloedy/Temporal-reconstruction-network)

## Authors’ contributions

Conceptualisations: AT, NH, Data curation: NL, Formal analysis: NL, AT, Methodology: NL, NH, AT, Supervision: NH, AT, Writing - original draft: NL; Writing - review & editing: JW, NH, AT. All authors contributed and reviewed the manuscript, and approved the final version for publication.

## Conflict of interest declaration

The authors declare no competing interests

## Funding

This work received funding from the European Research Council (ERC) under the European Union’s Horizon 2020 research and innovation programme (Grant No. 101003688 [EpiPose] and No. 101095619 [ESCAPE]). This work reflects only the authors’ view. The European Commission is not responsible for any use that may be made of the information it contains.

## Notes

### Competing Interest Statement

The authors have declared no competing interest.

### Funding Statement

This work received funding from the European Research Council (ERC) under the European Unions Horizon 2020 research and innovation programme (Grant No. 101003688 [EpiPose] and No. 101095619 [ESCAPE]). This work reflects only the authors view. The European Commission is not responsible for any use that may be made of the information it contains.

## References

Jones RM, Brosseau LM. 2015 Aerosol transmission of infectious disease. J. Occup. Environ. Med. 57, 501–508.

Loedy N et al. 2023 Longitudinal social contact data analysis: insights from 2 years of data collection in Belgium during the COVID-19 pandemic. BMC Public Health 23, 1298. (doi:10.1186/s12889-023-16193-7)

Beutels P, Shkedy Z, Aerts M, Van Damme P. 2006 Social mixing patterns for transmission models of close contact infections: exploring self-evaluation and diary-based data collection through a web-based interface. Epidemiol. Infect. 134, 1158–1166.

Hoang T, Coletti P, Melegaro A, Wallinga J, Grijalva CG, Edmunds JW, Beutels P, Hens N. 2019 A systematic review of social contact surveys to inform transmission models of close-contact infections. Epidemiol. Camb. Mass 30, 723. (doi:10.1097%2FEDE.0000000000001047)

Mossong J et al. 2008 Social contacts and mixing patterns relevant to the spread of infectious diseases. PLoS Med. 5, e74. (doi:10.1371/journal.pmed.0050074)

Wambua J et al. 2023 The influence of COVID-19 risk perception and vaccination status on the number of social contacts across Europe: insights from the CoMix study. BMC Public Health 23, 1350.

Gimma A et al. 2022 Changes in social contacts in England during the COVID-19 pandemic between March 2020 and March 2021 as measured by the CoMix survey: A repeated cross-sectional study. PLoS Med. 19, e1003907.

Jarvis CI, Van Zandvoort K, Gimma A, Prem K, Klepac P, Rubin GJ, Edmunds WJ. 2020 Quantifying the impact of physical distance measures on the transmission of COVID-19 in the UK. BMC Med. 18, 1–10.

Wong KL, Gimma A, Coletti P, Faes C, Beutels P, Hens N, Jaeger VK, Karch A, Johnson H. 2023 Social contact patterns during the COVID-19 pandemic in 21 European countries–evidence from a two-year study. BMC Infect. Dis. 23, 268.

Coletti P et al. 2020 CoMix: comparing mixing patterns in the Belgian population during and after lockdown. Sci. Rep. 10, 21885.

Tizzani M et al. 2023 Impact of tiered measures on social contact and mixing patterns of in Italy during the second wave of COVID-19. BMC Public Health 23, 906.

Goeyvaerts N, Santermans E, Potter G, Torneri A, Van Kerckhove K, Willem L, Aerts M, Beutels P, Hens N. 2018 Household members do not contact each other at random: implications for infectious disease modelling. Proc. R. Soc. B 285, 20182201.

Willem L, Verelst F, Bilcke J, Hens N, Beutels P. 2017 Lessons from a decade of individual-based models for infectious disease transmission: a systematic review (2006-2015). BMC Infect. Dis. 17, 1–16.

Abrams S et al. 2021 Modelling the early phase of the Belgian COVID-19 epidemic using a stochastic compartmental model and studying its implied future trajectories. Epidemics 35, 100449.

Willem L et al. 2021 The impact of contact tracing and household bubbles on deconfinement strategies for COVID-19. Nat. Commun. 12, 1524.

Coletti P et al. 2021 A data-driven metapopulation model for the Belgian COVID-19 epidemic: assessing the impact of lockdown and exit strategies. BMC Infect. Dis. 21, 1–12.

Held L, Hens N D O’Neill P, Wallinga J. 2019 Handbook of infectious disease data analysis. CRC Press.

Del Valle SY, Hyman JM, Hethcote HW, Eubank SG. 2007 Mixing patterns between age groups in social networks. Soc. Netw. 29, 539–554.

Wallinga J, Teunis P, Kretzschmar M. 2006 Using data on social contacts to estimate age-specific transmission parameters for respiratory-spread infectious agents. Am. J. Epidemiol. 164, 936–944. (doi:10.1093/aje/kwj317)

Eames KT. 2008 Modelling disease spread through random and regular contacts in clustered populations. Theor. Popul. Biol. 73, 104–111. (doi:10.1016/j.tpb.2007.09.007)

Smieszek T, Fiebig L, Scholz RW. 2009 Models of epidemics: when contact repetition and clustering should be included. Theor. Biol. Med. Model. 6, 1–15. (doi:10.1186/1742-4682-6-11)

Pung R, Firth JA, Russell T, Rogers T, Lee VJ, Kucharski AJ. 2023 Temporal contact patterns and the implications for predicting superspreaders and planning of targeted outbreak control. medRxiv, 2023–11.

Joe H, Zhu R. 2005 Generalized Poisson distribution: the property of mixture of Poisson and comparison with negative binomial distribution. Biom. J. J. Math. Methods Biosci. 47, 219–229.

Consul P, Famoye F. 1992 Generalized Poisson regression model. Commun. Stat.-Theory Methods 21, 89–109.

Britton T. 2010 Stochastic epidemic models: a survey. Math. Biosci. 225, 24–35. (doi:10.1016/j.mbs.2010.01.006)

Anderson RM, May RM. 1991 Infectious diseases of humans: dynamics and control. Oxford university press.

Biggerstaff M, Cauchemez S, Reed C, Gambhir M, Finelli L. 2014 Estimates of the reproduction number for seasonal, pandemic, and zoonotic influenza: a systematic review of the literature. BMC Infect. Dis. 14, 1–20. (doi:10.1186/1471-2334-14-480)

Achaiah NC, Subbarajasetty SB, Shetty RM. 2020 R0 and re of COVID-19: can we predict when the pandemic outbreak will be contained? Indian J. Crit. Care Med. Peer-Rev. Off. Publ. Indian Soc. Crit. Care Med. 24, 1125. (doi:10.5005%2Fjp-journals-10071-23649)

Munasinghe L, Asai Y, Nishiura H. 2019 Quantifying heterogeneous contact patterns in Japan: A social contact survey. Theor. Biol. Med. Model. 16, 1–10.

Prem K, Zandvoort K van, Klepac P, Eggo RM, Davies NG, Centre for the Mathematical Modelling of Infectious Diseases COVID-19 Working Group, Cook AR, Jit M. 2021 Projecting contact matrices in 177 geographical regions: an update and comparison with empirical data for the COVID-19 era. PLoS Comput. Biol. 17, e1009098. (doi:10.1371/journal.pcbi.1009098)

Kremer C, Torneri A, Boesmans S, Meuwissen H, Verdonschot S, Vanden Driessche K, Althaus CL, Faes C, Hens N. 2021 Quantifying superspreading for COVID-19 using Poisson mixture distributions. Sci. Rep. 11, 14107. (doi:10.1038/s41598-021-93578-x)

Stein RA. 2011 Super-spreaders in infectious diseases. Int. J. Infect. Dis. 15, e510–e513.

Duerr H-P, Schwehm M, Leary C, De Vlas S, Eichner M. 2007 The impact of contact structure on infectious disease control: influenza and antiviral agents. Epidemiol. Infect. 135, 1124–1132.

