## Supplementary Material for "Repetition in Social Contact Interactions: Implications in Modelling the Transmission of Respiratory Infectious Diseases in Pre-pandemic & Pandemic Settings"

Table of Contents:

**1. Summary of social contact studies** **2**
**2. Supplementary materials (Figures)** **4**
**3. Supplementary materials (Tables)** **14**
**4. Additional information** **16**

### 1. Summary of social contact studies

The POLYMOD study is a diary-based survey that investigated social contacts in eight countries: Belgium (BE), Germany (DE), Finland (FI), the United Kingdom (GB), Italy (IT), Luxembourg (LU), the Netherlands (NL), and Poland (PL) during the period of 2005-2006 [1]. This study was designed to cover different age categories and is a pioneer of large-scale prospectively collected, population-based surveys of epidemiologically relevant social contact patterns.

A similar survey, known as the CoMix study [2,3], was undertaken within the context of the COVID-19 pandemic. The primary objective of this study was to monitor public awareness and behaviour and was carried out in multiple countries. In both surveys, participants were asked to provide at least basic sociodemographic information and report contacts made on a day between 5 a.m. on the day preceding the survey day and 5 a.m. of the survey day. A contact was defined as either skin-to-skin contact such as a kiss or handshake (a physical contact), or a two-way conversation with three or more words in the physical presence of another person but no skin-to-skin contact (a nonphysical contact). Additionally, they were asked to provide supplementary information about these contacts. This included details such as age and the gender of the contacted person, whether the contact included skin-to-skin touching (physical/non-physical contact), the duration of the contact (less than 5 minutes, 5–15 minutes, 15 minutes to 1 hour, 1–4 hours, or 4 hours or more), the frequency (daily, weekly, monthly, a few times a year, or for the first time) with which they usually contact this person, and location of contacts (home, work, school, leisure activities, other places). A contact was defined as either an in-person conversation of three or more words in the physical presence of another person (such as a kiss or handshake) or without skin-to-skin contact.

Unlike the POLYMOD study, which is a cross-sectional survey relying on diaries or face-to-face interviews with participants, CoMix employs a bi-weekly online longitudinal survey. Consequently, the study is susceptible to participant fatigue, a phenomenon that may occur when participants become bored or less interested in the survey. In this study, we address potential under-reporting in the number of contacts resulting from this phenomenon [2]. We also extended this method to account for each participant's contact frequency distribution after correcting for under-reporting. We take into account participants' age, household size, area of residence, wave of participation, vaccination status, and microscopic time variables and their interactions to build Generalised Additive Models for Location, Scale, and Shape models with participants' ID as random intercepts. The comparison between the total number of contacts in the Belgian CoMix study and those obtained by correcting the under-reporting due to fatigue is presented in **Figure S1**.

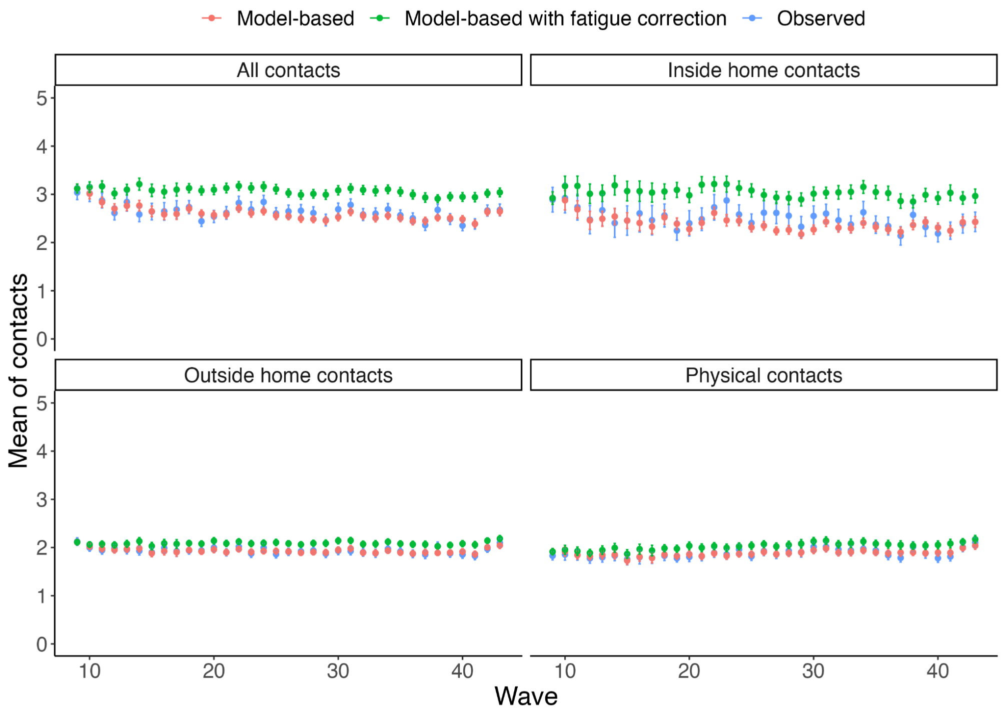


**Figure S1.** Comparison of the total number of contacts from the Belgian CoMix study between observed, mode-based, and model-based with under-reporting due to fatigue correction in different settings.

### 2. Supplementary materials (Figures)

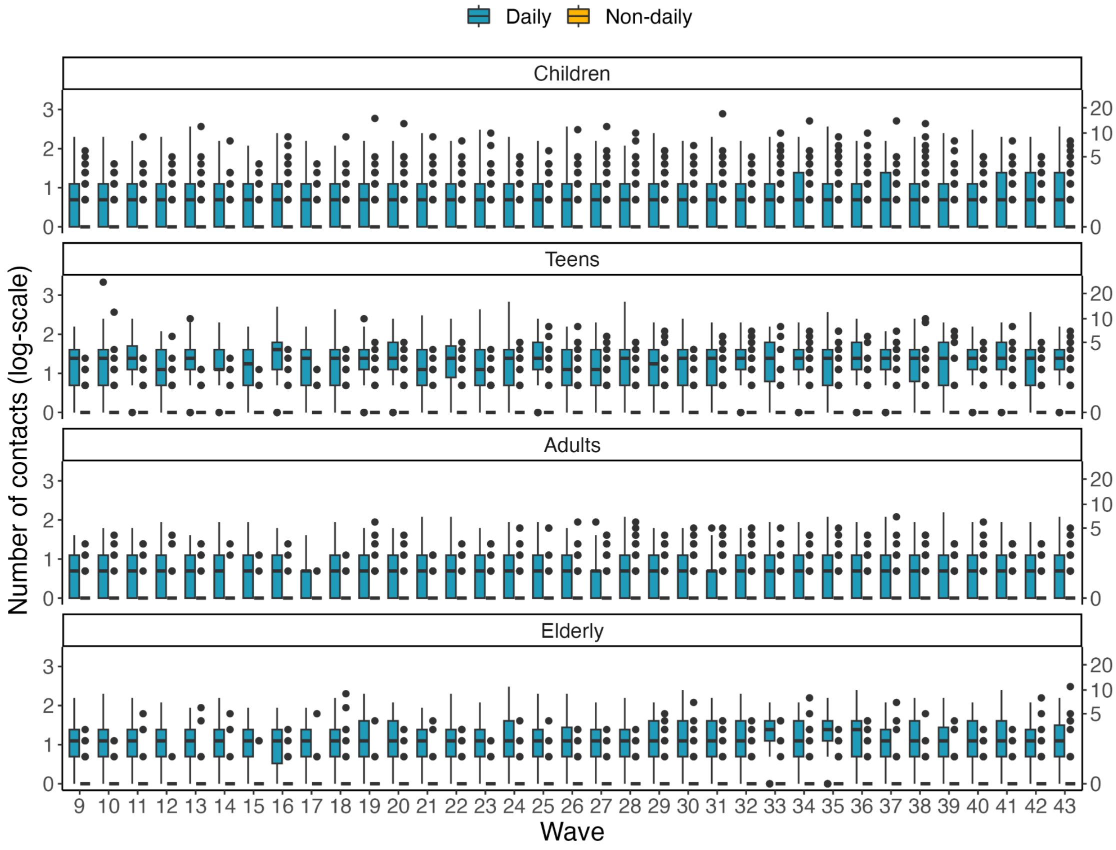


**Figure S2.** The distribution of the total number of daily and non-daily physical contacts from the CoMix study.

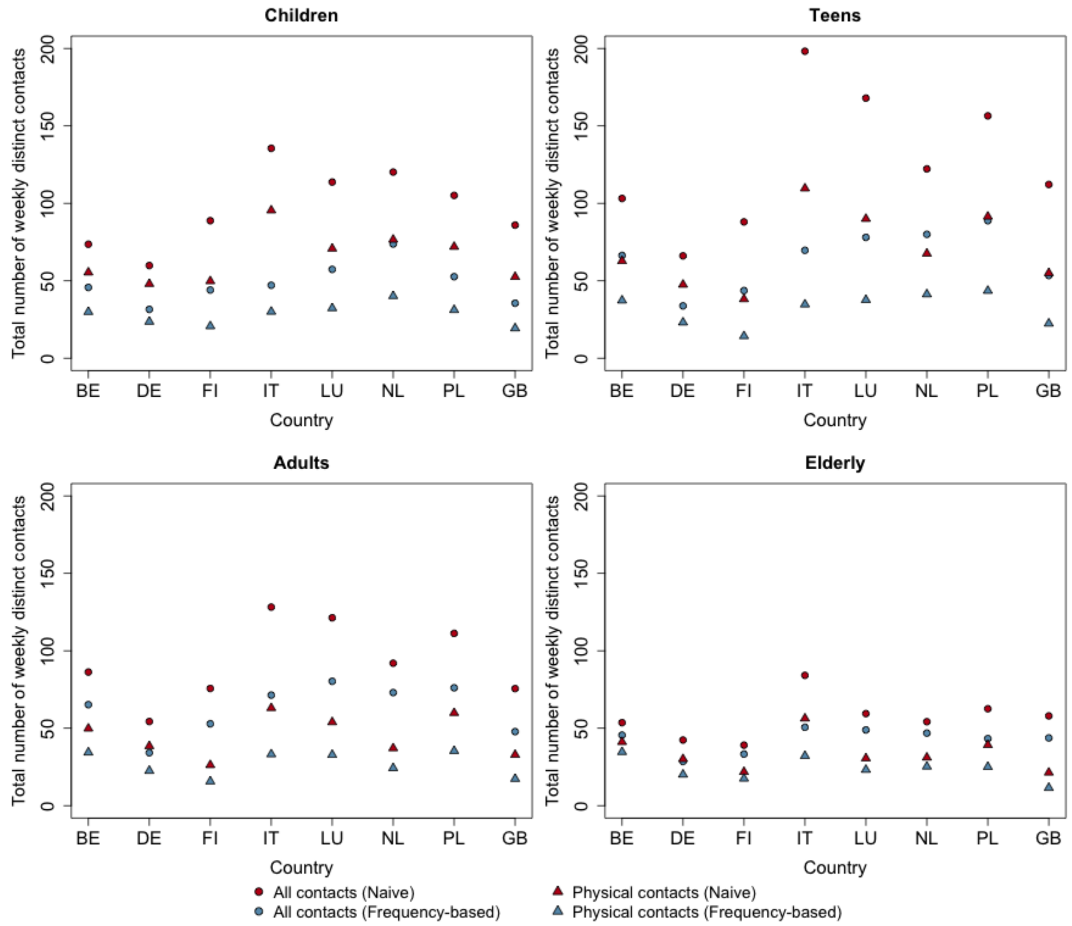


**Figure S3.** Comparison between the number of distinct contacts in a week calculated via the frequency-based and naive approach for the pre-pandemic scenario.

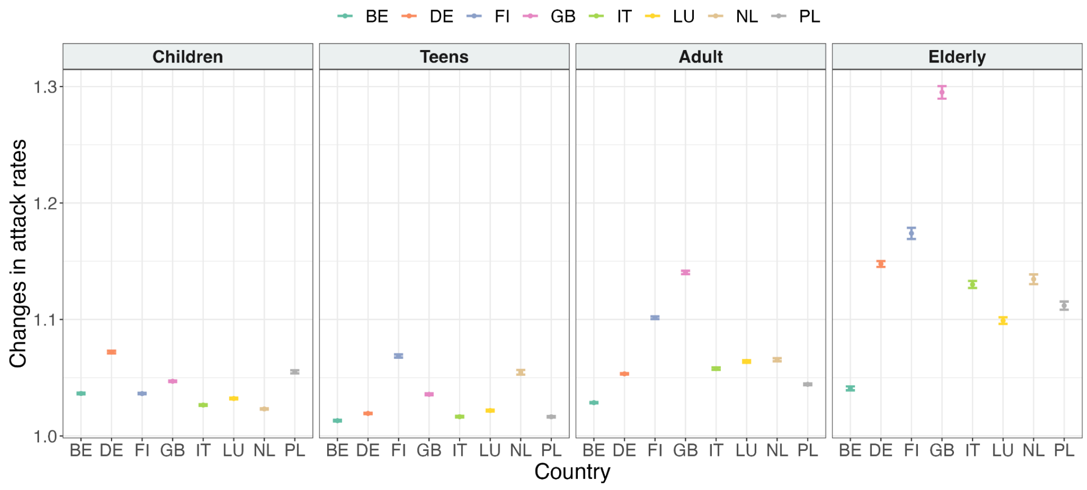


**Figure S4.** Changes in epidemic attack rates in COVID-19-like illness when simulating epidemics using the frequency-based approach compared to the naive approach during a pre-pandemic scenario, along with non-parametric bootstrapping for 95% confidence intervals.

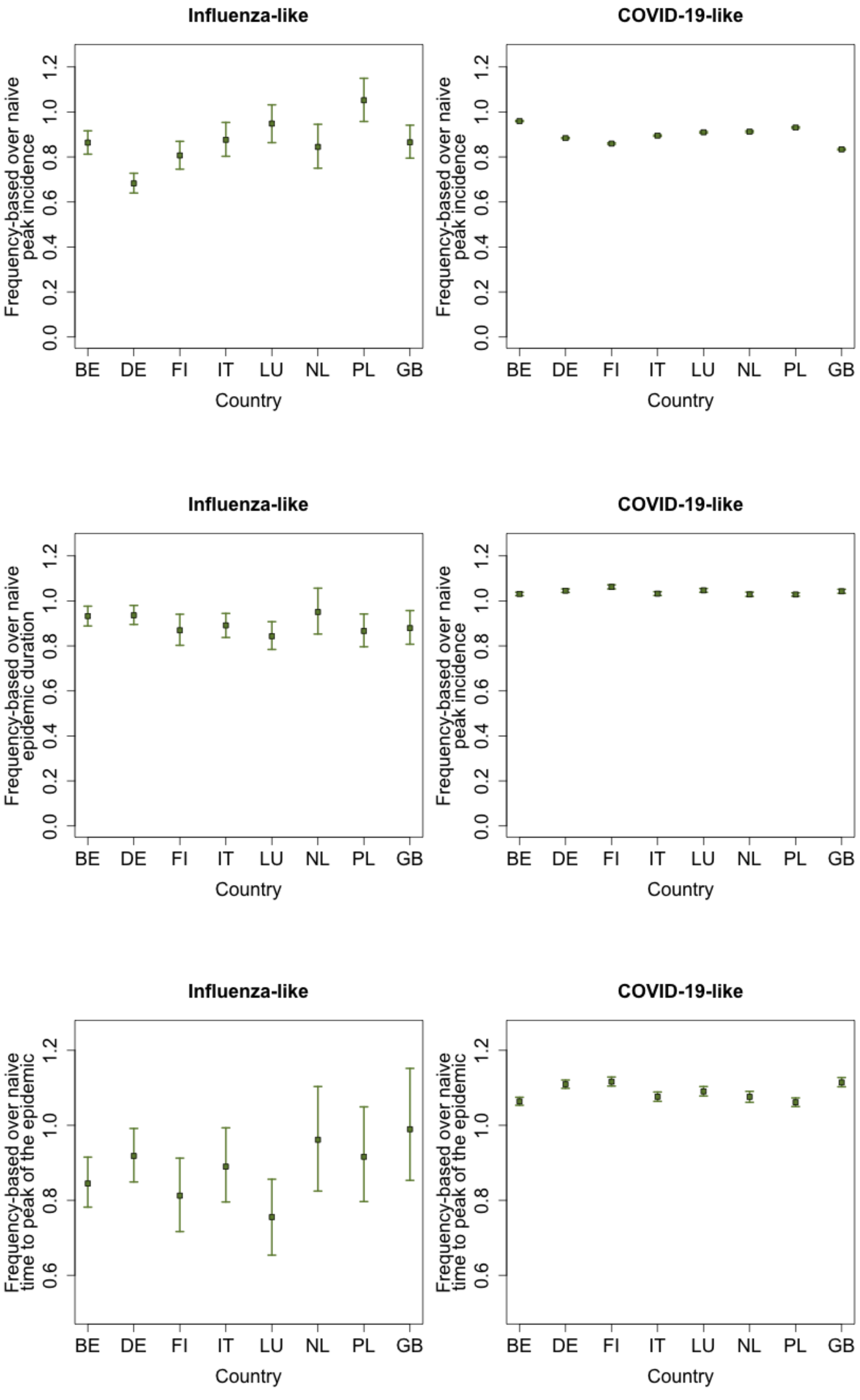
Influenza-like
COVID-19-like
Frequency-based over naive peak incidence
Frequency-based over naive epidemic duration
Frequency-based over naive time to peak of the epidemic
0.0
0.2
0.4
0.6
0.8
1.0
1.2
BE
DE
FI
IT
LU
NL
PL
GB
Country

**Figure S5.** The ratio between frequency-based and naive approach epidemic simulations for the pre-pandemic setting, illustrating outcomes for peak incidence (Top), epidemic duration (Middle), and time to peak of the epidemic (Bottom), along with the 95% Confidence Intervals derived from non-parametric bootstrap.

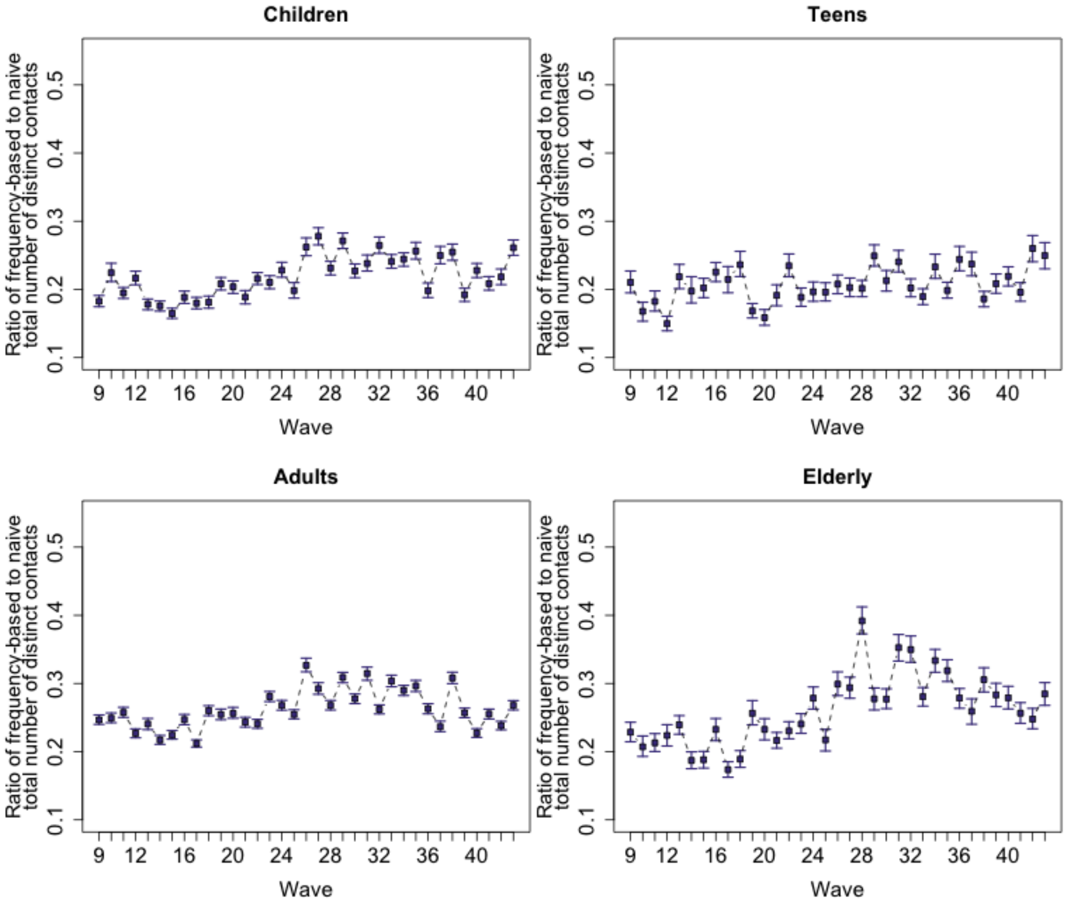


**Figure S6.** The ratio between the frequency-based and naively calculated distinct physical contacts in a week for Children, Teens, Adults, and the Elderly in the pandemic setting with the 95% Confidence Intervals from the non-parametric bootstrap.

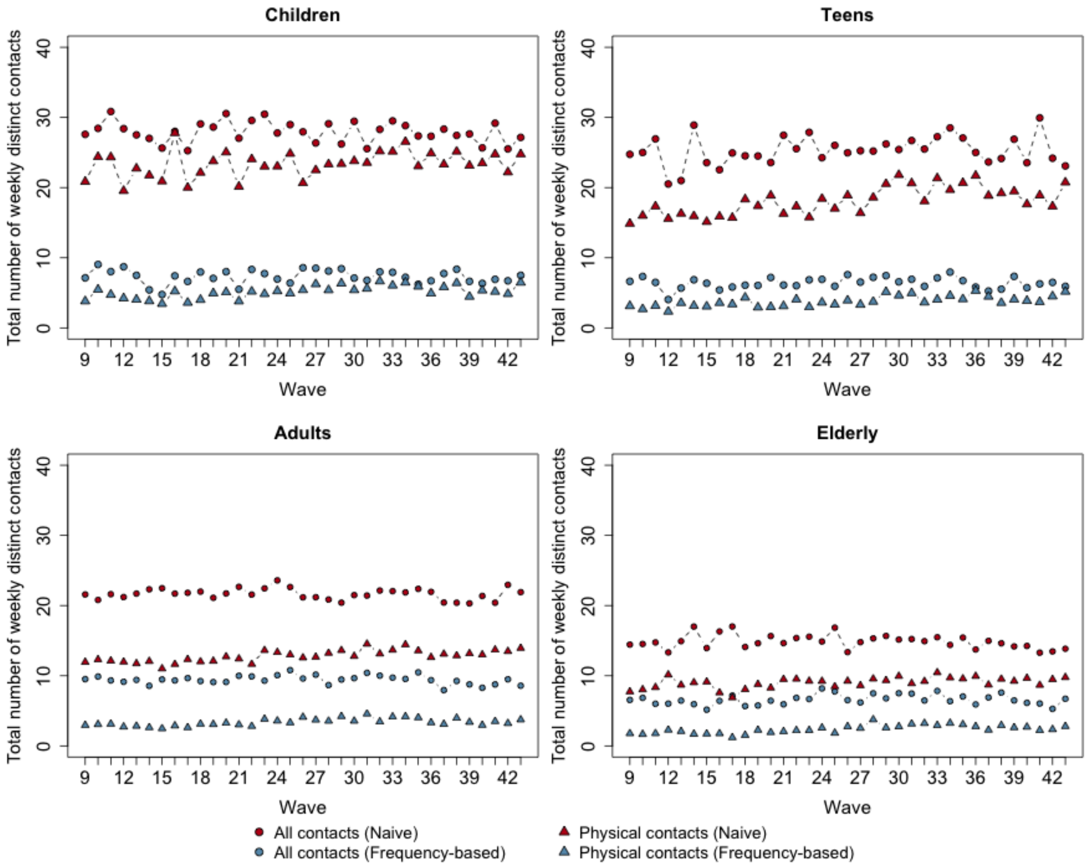


**Figure S7.** Comparison between the number of distinct contacts in a week calculated via the frequency-based and naive approach for the pandemic scenario.

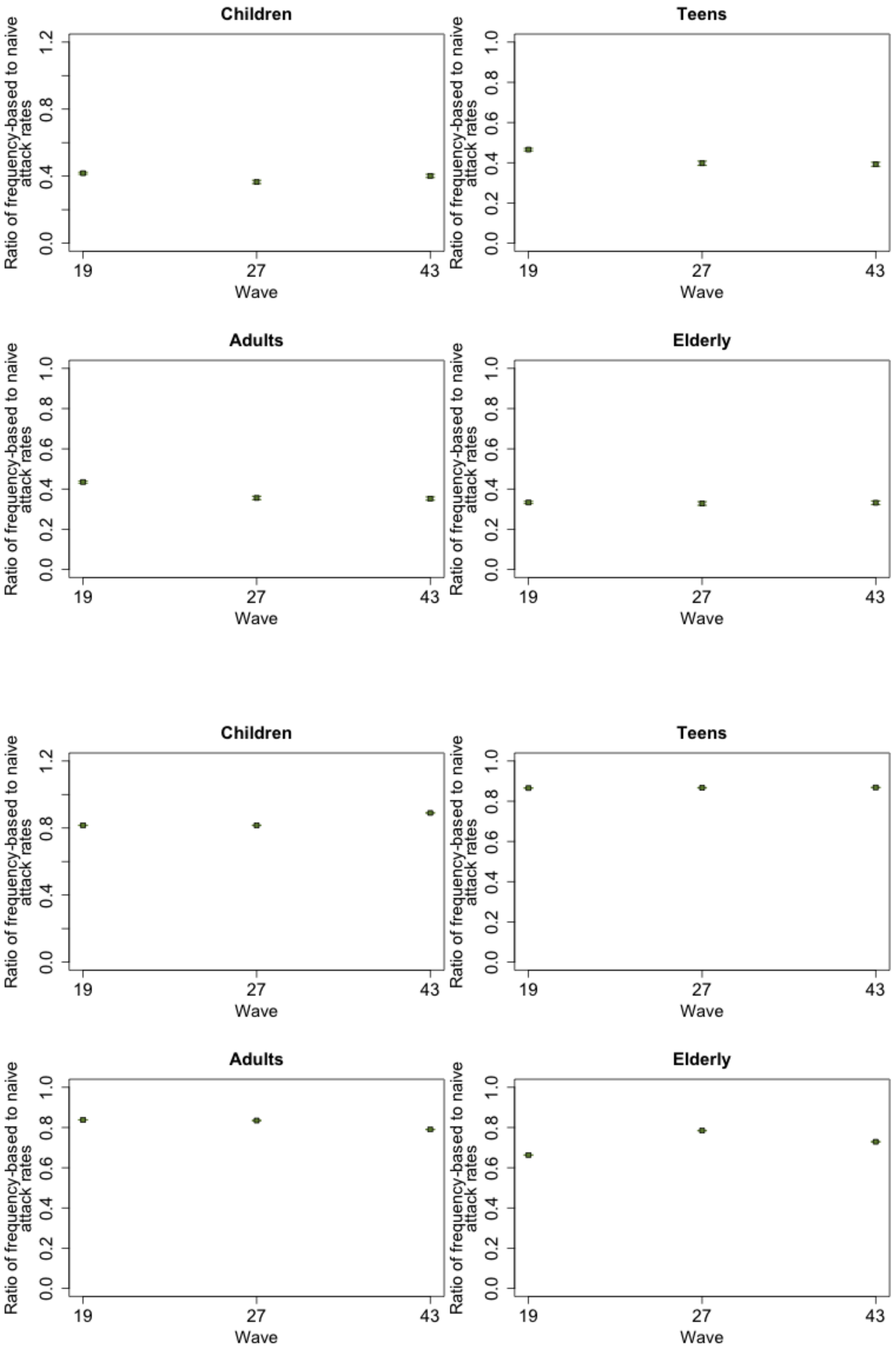


**Figure S8.** The Ratio of epidemics attack rate for the frequency-based approach in comparison with the naive approach with the physical contacts from the CoMix study, together with the non-parametric bootstrap for the 95% confidence intervals. *(Top)*

Influenza-like *(Bottom)* COVID-19-like.

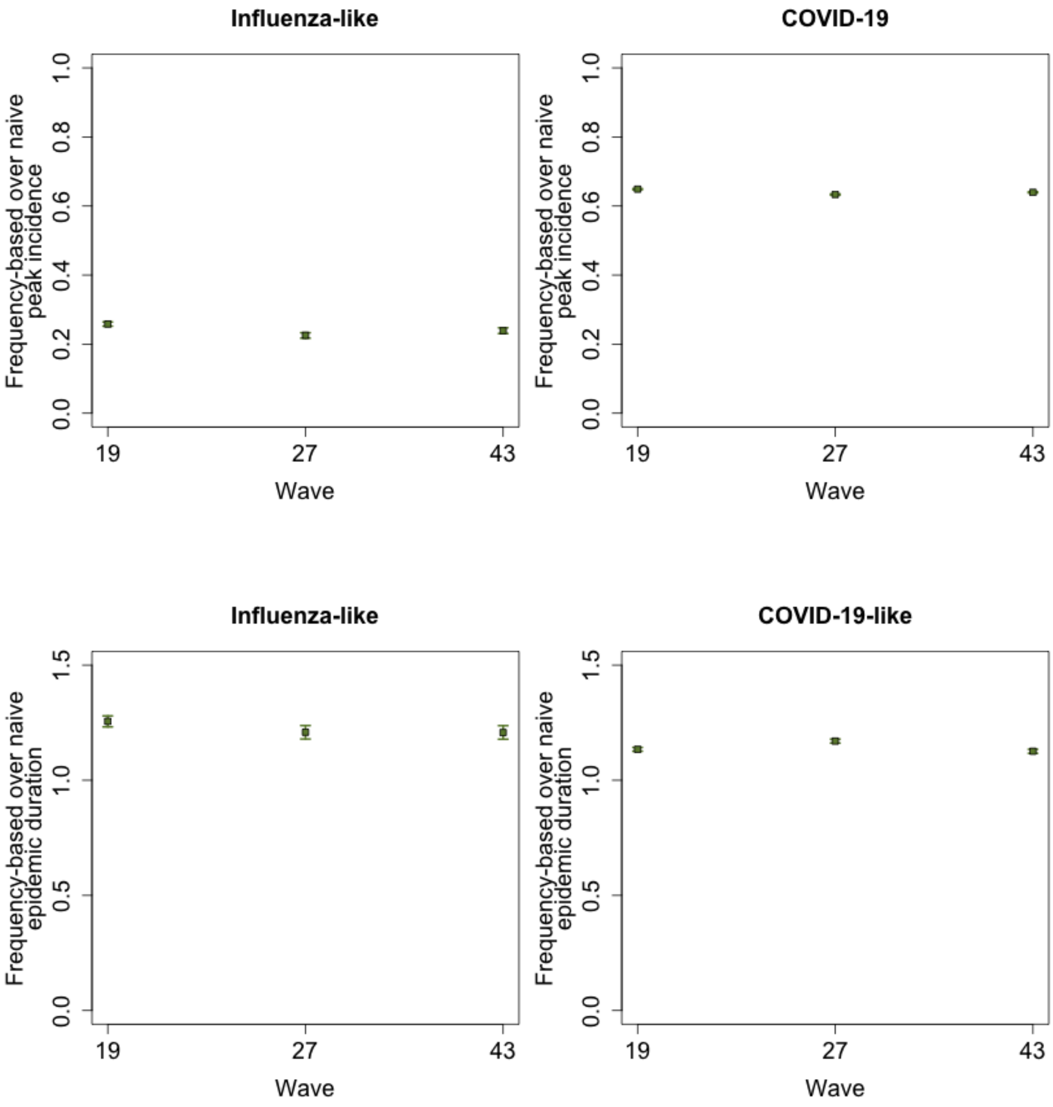

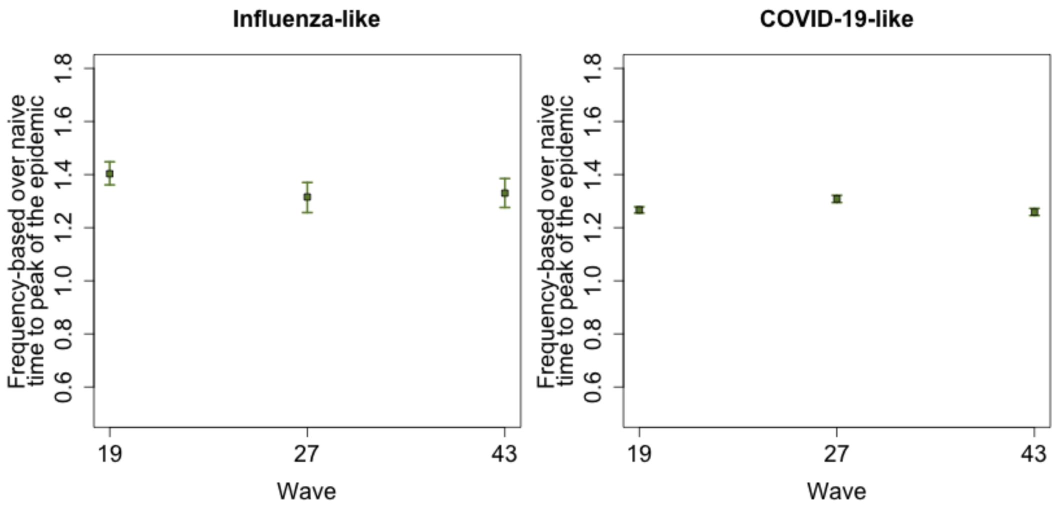


**Figure S9.** The ratio between frequency-based and naive approach epidemic simulations for the pandemic setting, illustrating outcomes for peak incidence (Top), epidemic duration (Middle), and time to peak of the epidemic (Bottom), along with the 95% Confidence Intervals derived from non-parametric bootstrap.

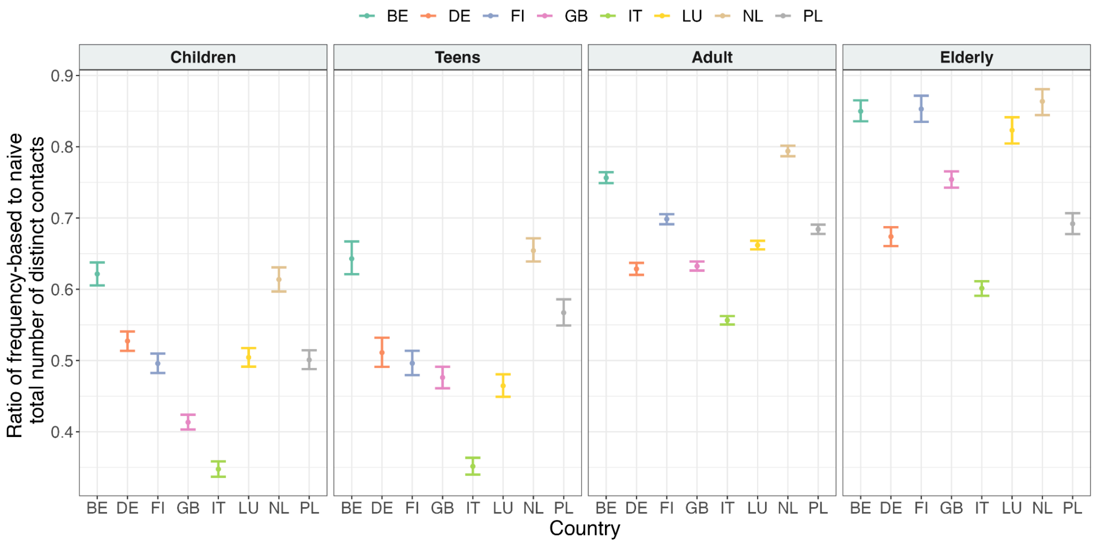


**Figure S10.** The ratio between frequency-based and naively calculated distinct contacts (all contacts) over a week in a pre-pandemic scenario, together with its 95% Confidence Interval obtained by a non-parametric bootstrap for Children, Teens, Adults, and the Elderly in different countries.

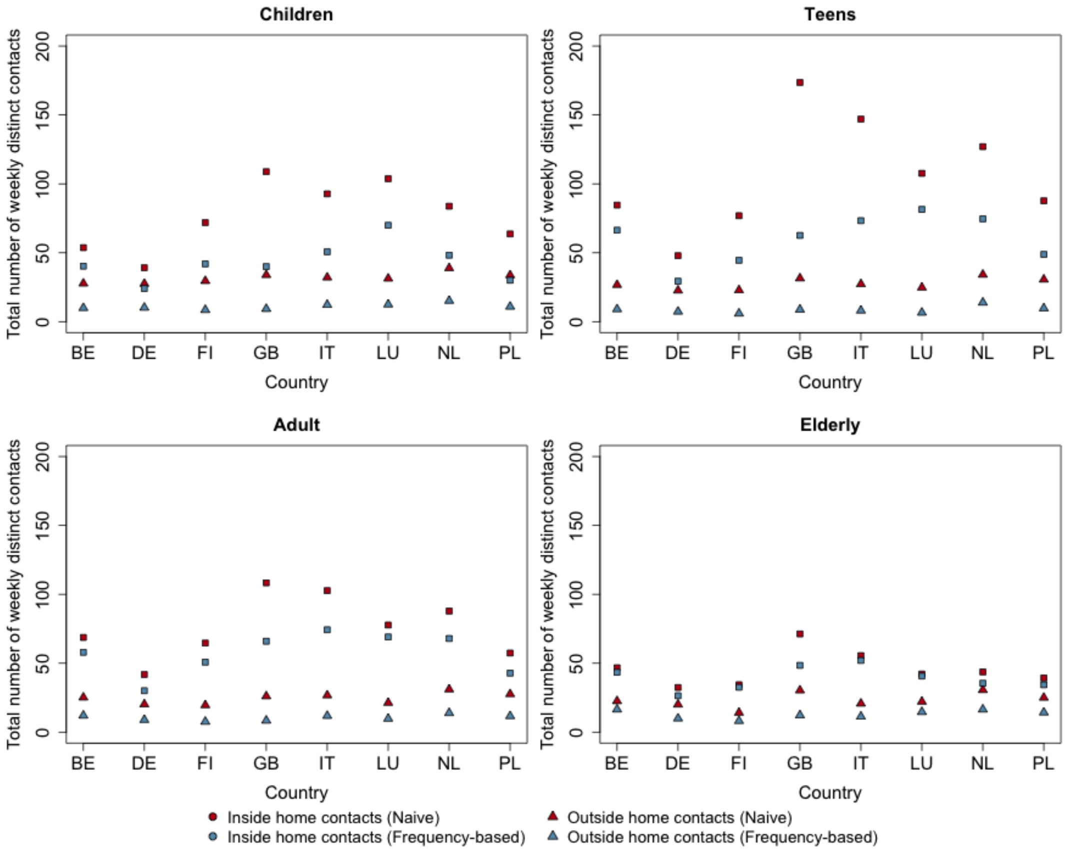


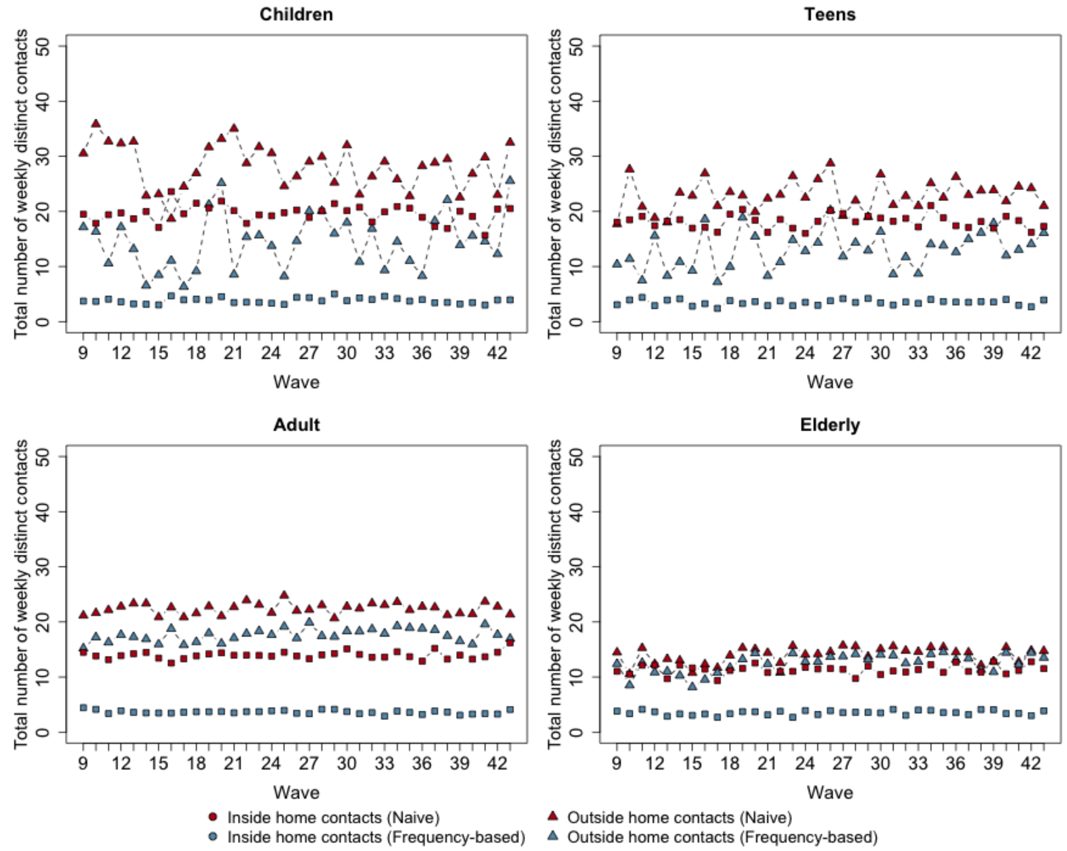


**Figure S11.** Comparison between the frequency-based approach and naive approach on the total number of weekly distinct contacts based on locations in *(Top)*

pre- and *(Bottom)* pandemic scenarios.

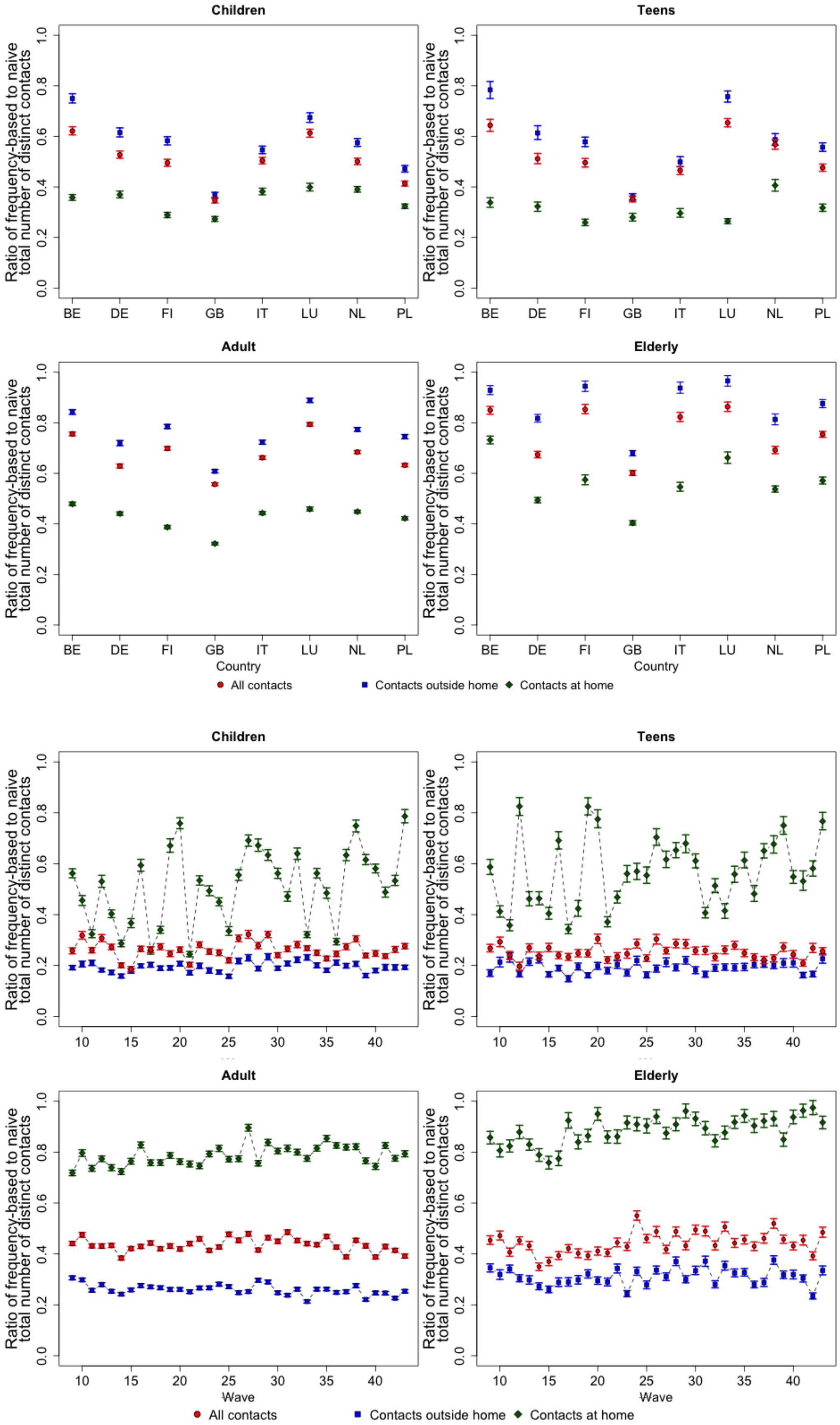


**Figure S12.** The ratio between frequency-based and naively calculated distinct physical contacts in a week based on the location of contacts for Children, Teens, Adults, and the Elderly in *(Top)* pre- and *(Bottom)* pandemic scenarios with the 95% Confidence Intervals from the non-parametric bootstrap.

### 3. Supplementary materials (Tables)

| | **Mon** | **Tue** | **Wed** | **Thu** | **Fri** | **Sat** | **Sun** |
|---|---|---|---|---|---|---|---|
| **Daily** | 4 | 4 | 4 | 4 | 4 | 4 | 4 |
| **Weekly** | 1 | 3 | 2 | 0 | 2 | 1 | 3 |
| **Monthly** | 1 | 0 | 1 | 2 | 2 | 0 | 3 |
| **A few times a year** | 1 | 1 | 0 | 3 | 0 | 0 | 0 |
| **First time** | 0 | 2 | 1 | 2 | 3 | 0 | 0 |

**Table S1.** Dummy result for the temporal reconstruction of an individual's contact pattern.

| | Influenza-like | | COVID-19-like | |
|---|---|---|---|---|
| | Baseline | Temporal | Baseline | Temporal |
| Belgium | 0.866 | 0.940 | 0.351 | 0.420 |
| Finland | 0.850 | 0.951 | 0.325 | 0.399 |
| Germany | 0.817 | 0.984 | 0.387 | 0.468 |
| Great Britain | 0.933 | 0.979 | 0.387 | 0.455 |
| Italy | 0.943 | 0.985 | 0.382 | 0.498 |
| Luxembourg | 0.950 | 0.991 | 0.446 | 0.516 |
| Netherlands | 0.952 | 0.985 | 0.377 | 0.449 |
| Poland | 0.911 | 0.986 | 0.393 | 0.486 |

**Table S2.** Comparison of the epidemics extinction rate obtained from the epidemics simulation using the temporal and baseline approaches with the POLYMOD study.

| | Influenza-like | | COVID-19-like | |
|---|---|---|---|---|
| | Baseline | Temporal | Baseline | Temporal |
| Wave 19 | 0.505 | 0.822 | 0.165 | 0.429 |
| Wave 27 | 0.555 | 0.880 | 0.168 | 0.431 |
| Wave 43 | 0.582 | 0.881 | 0.209 | 0.425 |

**Table S3.** Comparison of the epidemics extinction rate obtained from the epidemics simulation using the temporal and baseline approaches with the CoMix study.

| Infectious period | Age category | Influenza-like ($R_0$= 1.3) | COVID-19-like ($R_0$= 3.3) |
|---|---|---|---|
| 5 days | Children | 0.809 (0.753 - 0.866) | 0.960 (0.959 - 0.961) |
| | Teens | 0.861 (0.803 - 0.921) | 0.984 (0.983 - 0.985) |
| | Adults | 0.826 (0.770 - 0.884) | 0.968 (0.968 - 0.969) |
| | Elderly | 0.801 (0.733 - 0.868) | 0.956 (0.955 - 0.958) |
| 7 days | Children | 0.796 (0.760 - 0.834) | 0.965 (0.964 - 0.966) |
| | Teens | 0.833 (0.796 - 0.871) | 0.987 (0.986 - 0.988) |
| | Adults | 0.803 (0.768 - 0.841) | 0.972 (0.972 - 0.973) |
| | Elderly | 0.804 (0.761 - 0.846) | 0.961 (0.959 - 0.962) |
| 10 days | Children | 0.782 (0.757 - 0.808) | 0.968 (0.967 - 0.969) |
| | Teens | 0.823 (0.797 - 0.848) | 0.989 (0.988 - 0.989) |
| | Adults | 0.798 (0.774 - 0.822) | 0.975 (0.974 - 0.975) |
| | Elderly | 0.793 (0.766 - 0.819) | 0.965 (0.963 - 0.966) |

**Table S4.** The ratio of epidemic attack rates for the temporal approach in comparison with the baseline approach with the POLYMOD data for different infectious periods, together with the non-parametric bootstrap for the confidence intervals.

### 4. Additional information

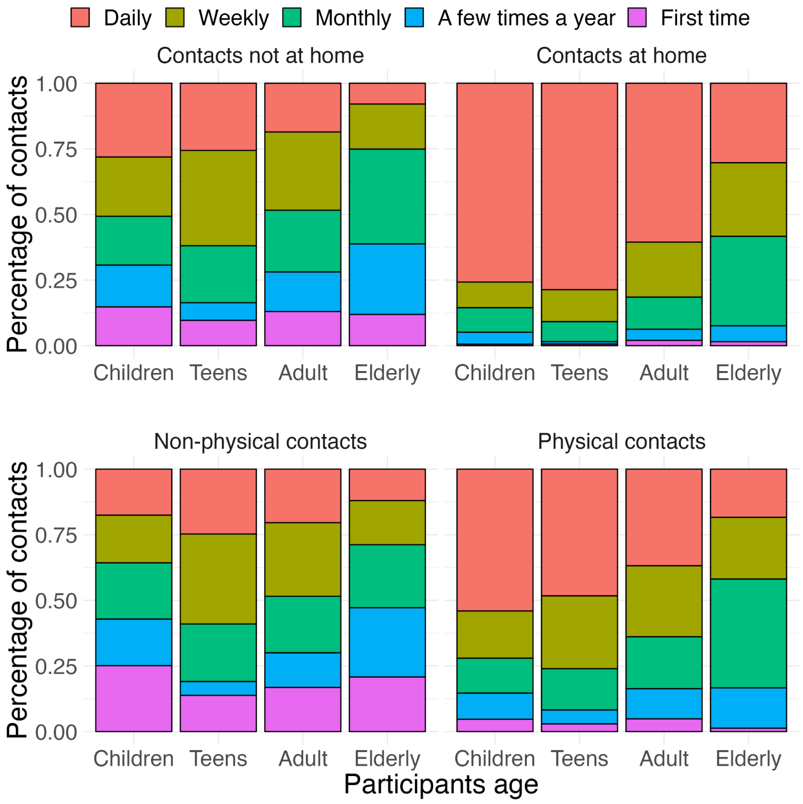


**Figure S13.** The proportion of contacts that occurred at home *(Top)* and contacts that involved physical contacts *(Bottom)* in Belgium

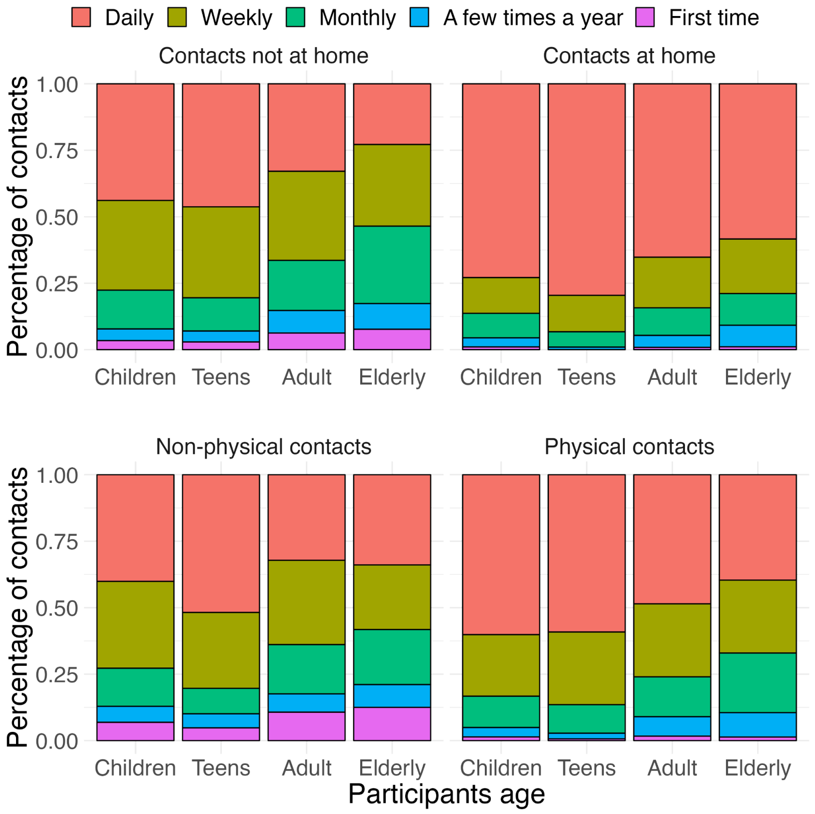


**Figure S14.** The proportion of contacts that occurred at home *(Top)* and contacts that involved physical contacts *(Bottom)* in Deutschland.

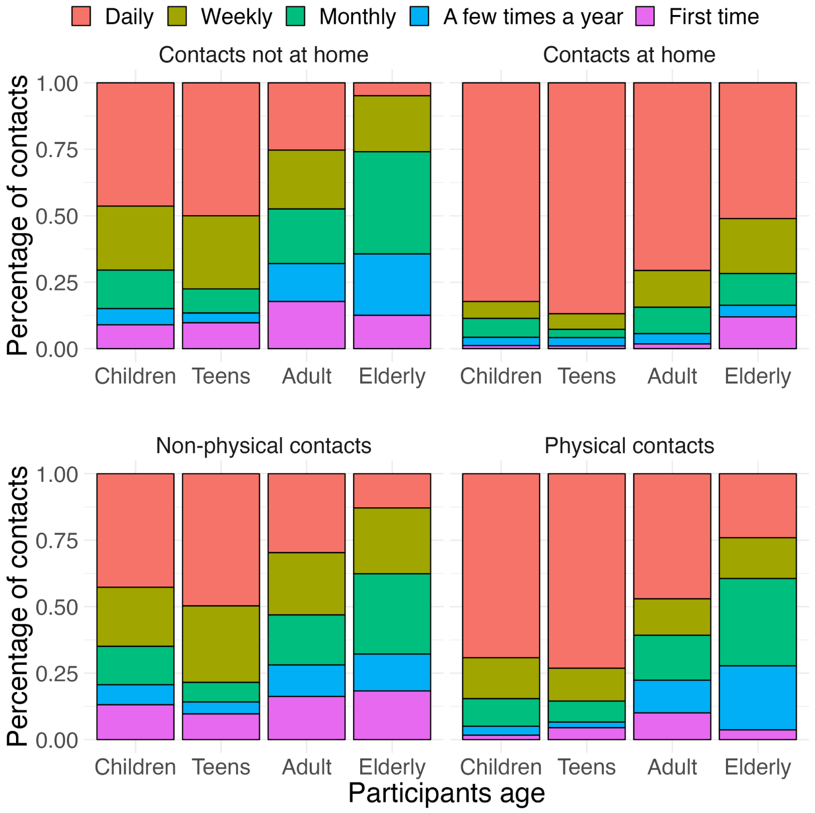


**Figure S15.** The proportion of contacts that occurred at home *(Top)* and contacts that involved physical contacts *(Bottom)* in Finland.

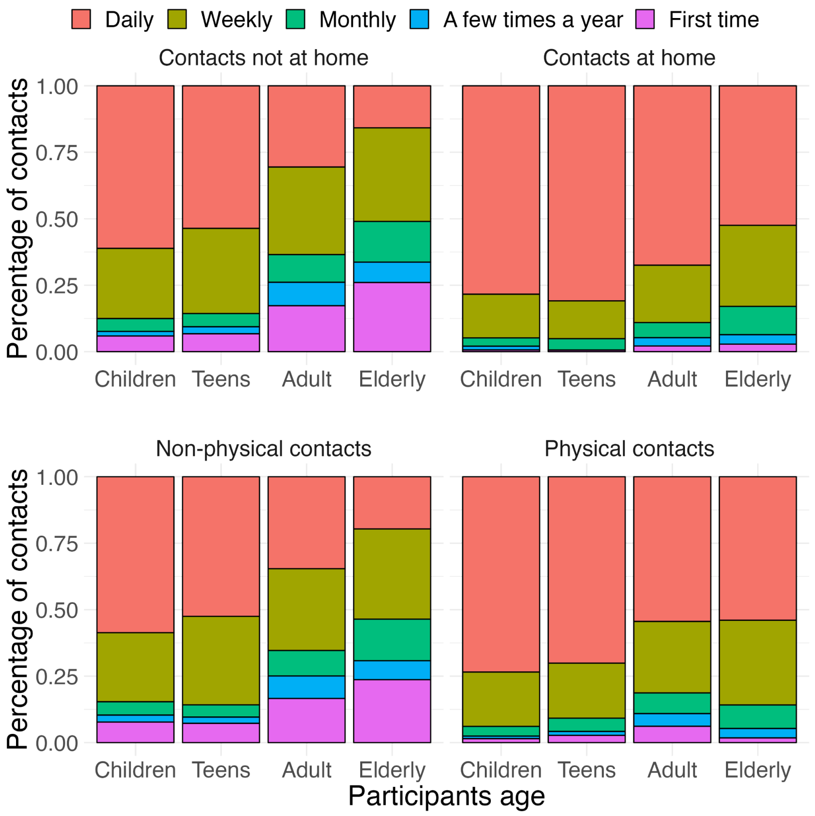


**Figure S16.** The proportion of contacts that occurred at home *(Top)* and contacts that involved physical contacts *(Bottom)* in Great Britain.

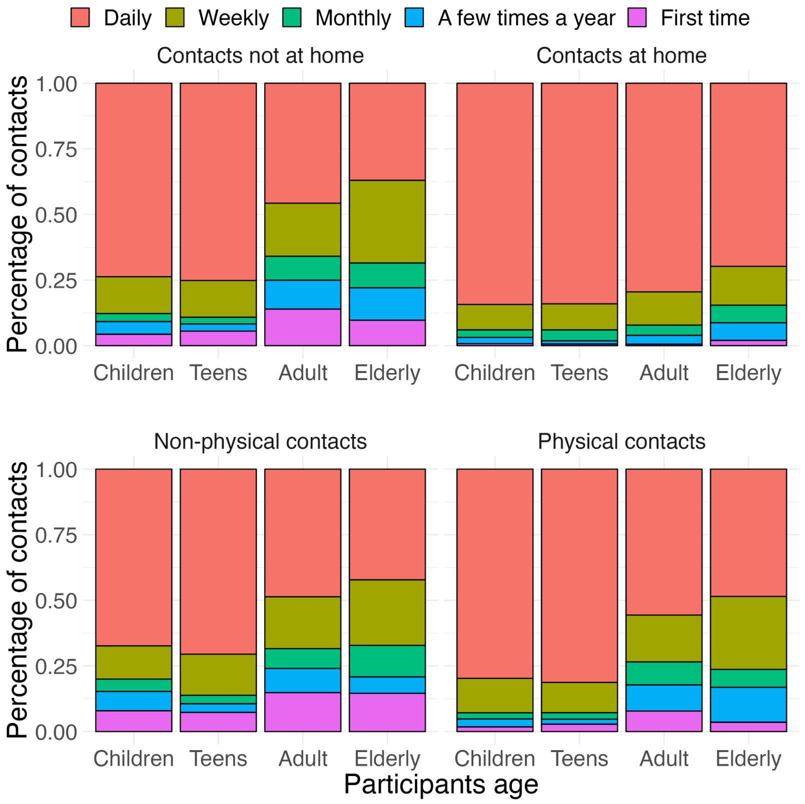


**Figure S17.** The proportion of contacts that occurred at home *(Top)* and contacts that involved physical contacts *(Bottom)* in Italy.

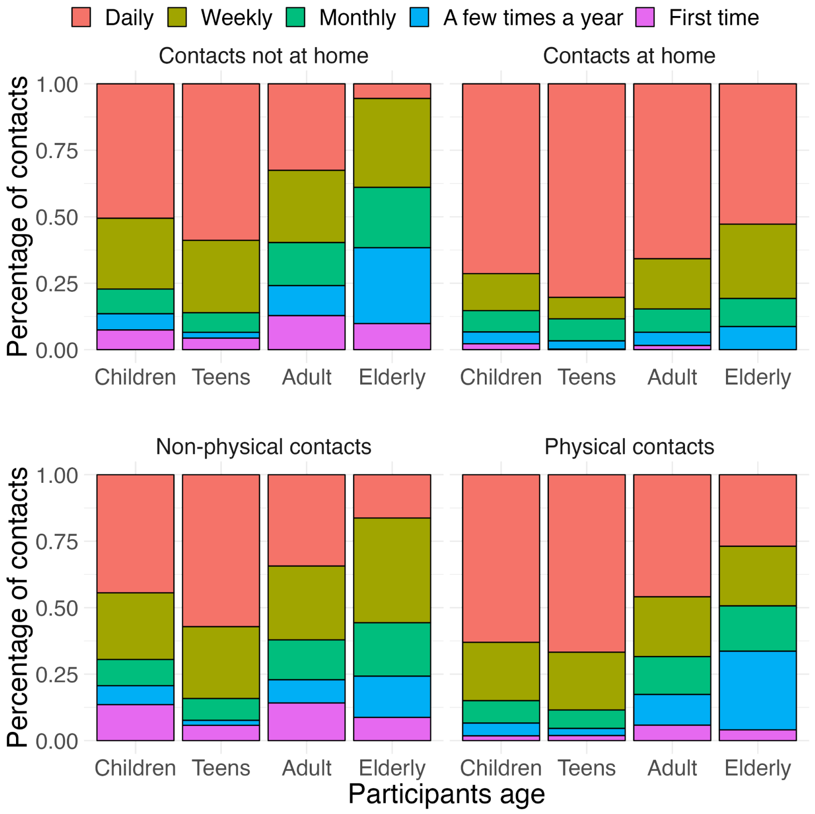


**Figure S18.** The proportion of contacts that occurred at home *(Top)* and contacts that involved physical contacts *(Bottom)* in Luxembourg.

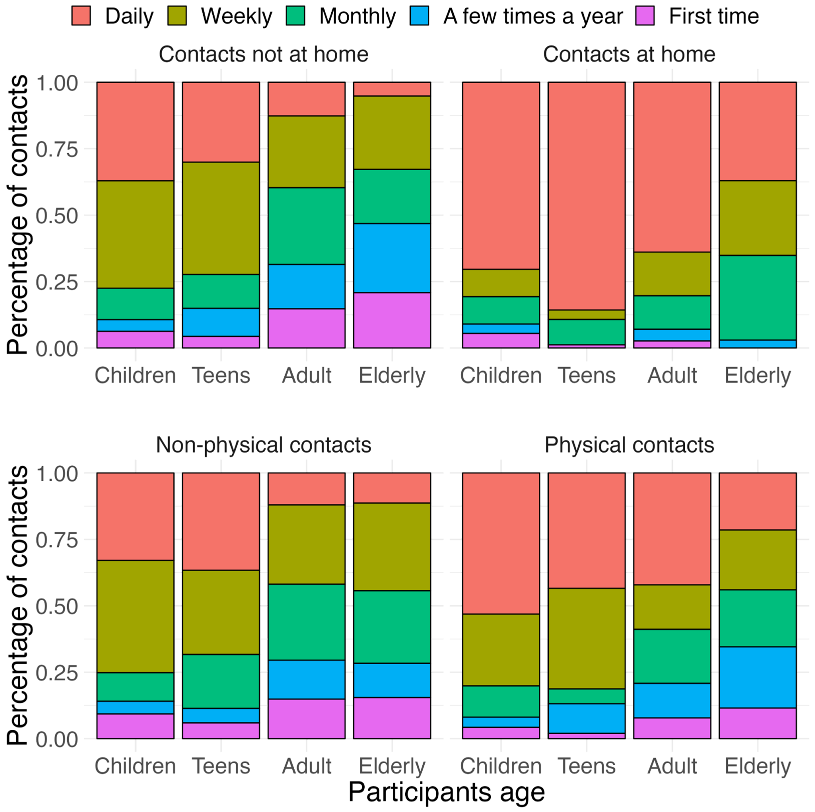


**Figure S19.** The proportion of contacts that occurred at home *(Top)* and contacts that involved physical contacts *(Bottom)* in the Netherlands.

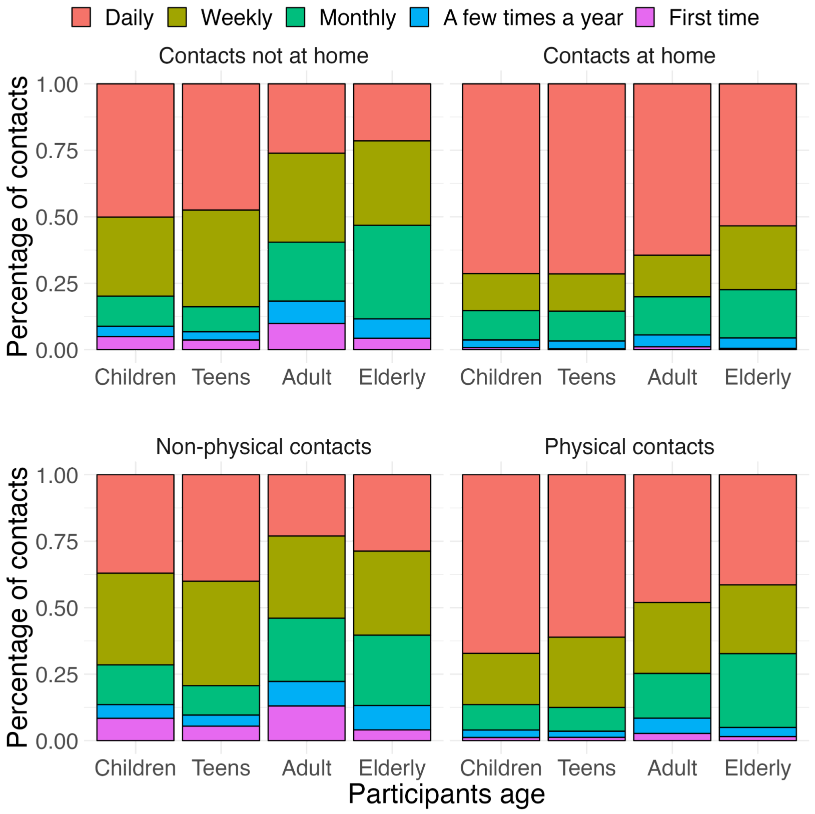


**Figure S20.** The proportion of contacts that occurred at home *(Top)* and contacts that involved physical contacts *(Bottom)* in Poland.

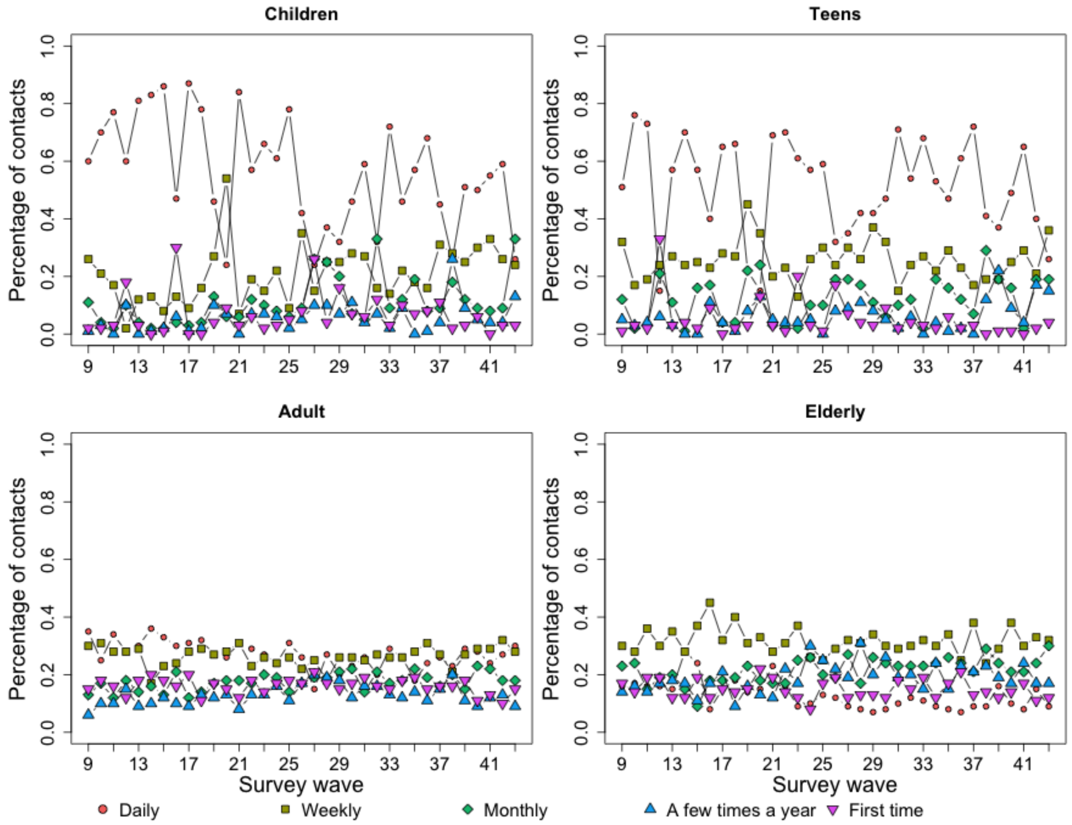


**Figure S21.** The proportion of contacts that occurred not at home in the CoMix study

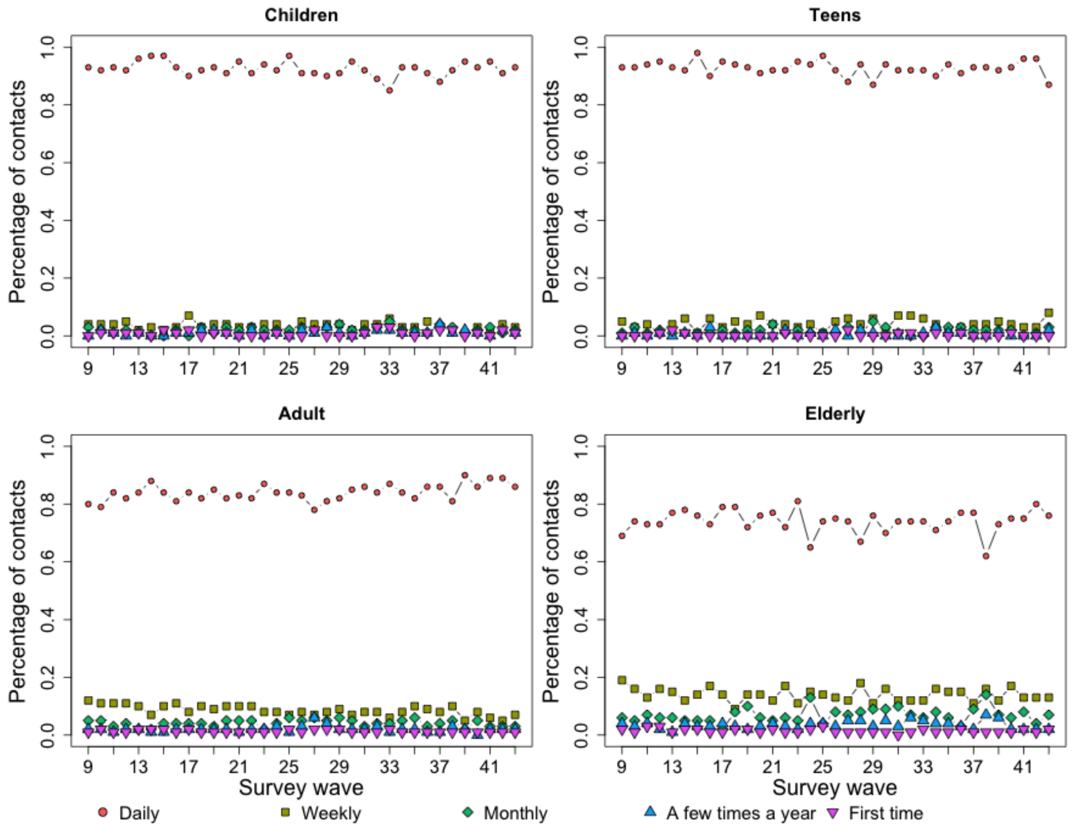


**Figure S22.** The proportion of contacts that occurred at home in the CoMix study

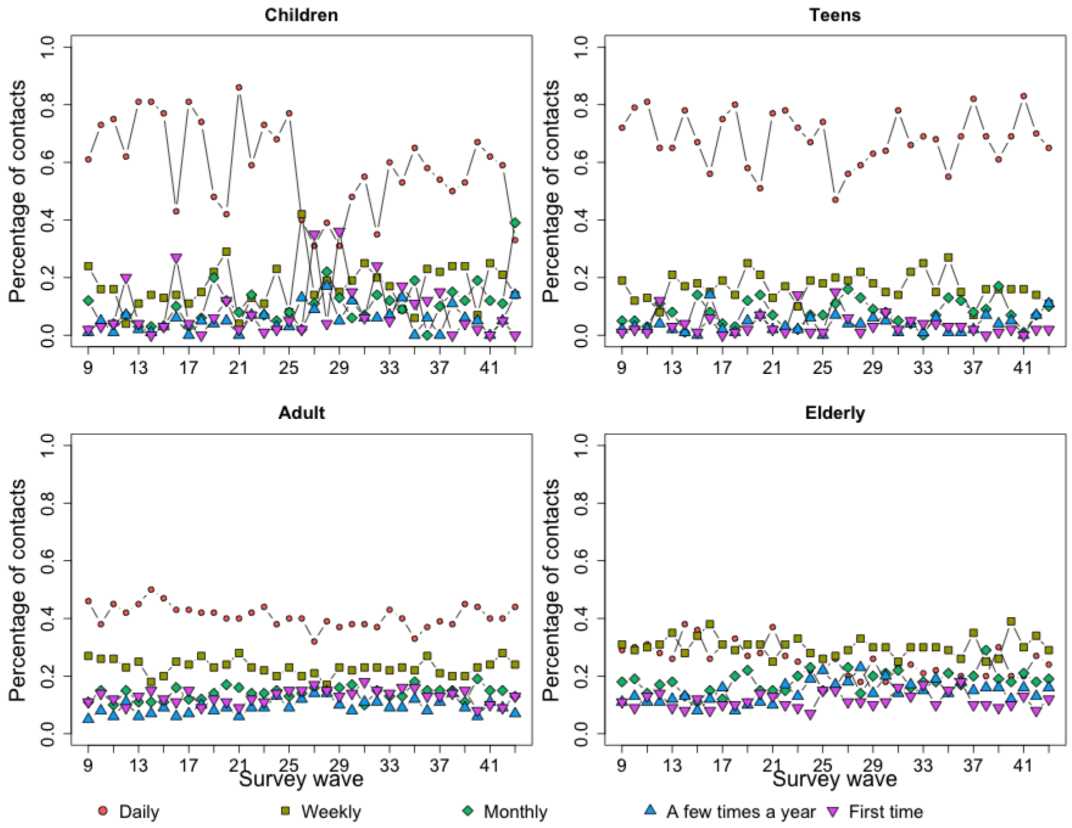


**Figure S23.** The proportion of non-physical contacts in the CoMix study

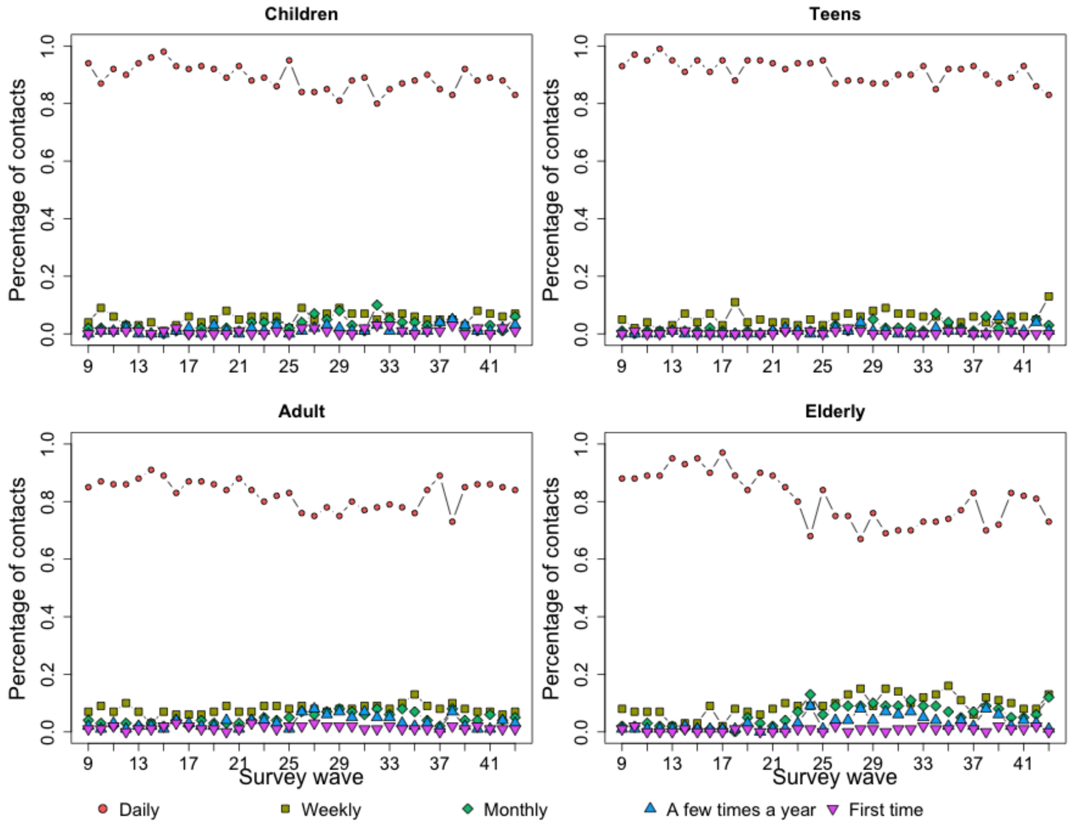


**Figure S24.** The proportion of physical contacts in the CoMix study

### References


1. Mossong J *et al.* 2008 Social contacts and mixing patterns relevant to the spread of infectious diseases. *PLoS Med.* **5**, e74. (doi:https://doi.org/10.1371/journal.pmed.0050074)
2. Loedy N *et al.* 2023 Longitudinal social contact data analysis: insights from 2 years of data collection in Belgium during the COVID-19 pandemic. *BMC Public Health* **23**, 1298. (doi:10.1186/s12889-023-16193-7)
3. Verelst F *et al.* 2021 SOCRATES-CoMix: a platform for timely and open-source contact mixing data during and in between COVID-19 surges and interventions in over 20 European countries. *BMC Med.* **19**, 1–7. (doi:https://doi.org/10.1186/s12916-021-02133-y)
